# DeepSpot-M: a multimodal foundation model for transcriptome-wide virtual spatial transcriptomics from histology

**DOI:** 10.64898/2026.06.19.26356060

**Authors:** Kalin Nonchev, Sebastian Dawo, Karina Silina, Viktor H Koelzer, Gunnar Rätsch

## Abstract

Spatial transcriptomics remains costly and low-throughput, limiting it to a small fraction of routine histology and leaving the molecular state of disease unmeasured in most patients. Predicting spatial expression from histology could address this gap, but existing methods are restricted to predefined genes and small cohorts. We present DeepSpot-M, a multimodal foundation model that predicts spatial expression by representing genes with embeddings from foundation models spanning DNA, RNA, proteins, single cells and biomedical text. By reformulating prediction as a query over genes, DeepSpot-M spans the protein-coding transcriptome and predicts genes unseen during training. Trained on a large pan-cancer dataset, it transfers to held-out cancers, outperforming specialised models trained on them, and adapts to new cohorts and single-cell assays from one slide via test-time adaptation. Applied to TCGA, it generates a virtual atlas of 28,664 slides across 32 cancers, recovering a pan-cancer map of malignancy from histology. The same query interface further enables transcriptome restoration, cross-species non-coding RNA inference, in silico variant-effect mapping and natural-language querying. We anticipate DeepSpot-M will provide a scalable foundation for virtual spatial transcriptomics and biomarker discovery.

## 1 Introduction

Routine histology is acquired for nearly every cancer patient, but the gene expression programmes that shape tissue morphology cannot be directly quantified from a stained slide [1–4]. Spatial transcriptomics (ST) measures gene expression in situ while preserving tissue architecture, enabling the study of cells in their native tissue context [5–8]. Yet current ST technologies remain costly, technically demanding and difficult to deploy at clinical scale [9, 10]. Computationally predicting spatial gene expression directly from routine histology could therefore provide a scalable route to molecular tissue characterisation, extending biomarker discovery to large retrospective cohorts, low-resource settings and, ultimately, routine diagnostic workflows [11, 12].

Recent deep learning approaches have demonstrated that spatial gene expression can be inferred from histological images [13–16]. Models ranging from convolutional networks to pathology foundation models can recover spatial expression patterns for subsets of genes from paired histology and ST data. For instance, recent approaches combine a pathology foundation model as a feature extractor with multi-scale spatial tissue context to infer spatial transcriptomic profiles [14]. However, two limitations remain [11, 12]. First, most models are trained on small, single-cohort datasets, which limits transfer across tissues, diseases and assays. Second, they treat genes as fixed output targets, restricting prediction to a predefined panel and preventing inference for genes not seen during training. Addressing both requires models trained on diverse cohorts that represent genes within a shared molecular space rather than as fixed outputs.

In fact, foundation models now provide shared molecular representations across biology, with each modality contributing a different view of a gene. Models of DNA capture regulatory syntax and sequence-function relationships [17–19], RNA models add the features of coding and non-coding transcripts, including isoform diversity [20], and protein models infer structural and functional properties from sequence [21, 22]. Complementary to these sequence-based views, single-cell models learn cellular states and gene co-expression programmes [23, 24], while biomedical language models distil gene-disease associations from the literature [25–27]. Individually, these models encode transferable biological priors, but they remain largely unimodal and disconnected from tissue morphology and spatial context. We reasoned that their gene-level embeddings could together provide a shared description of molecular identity on which to build histology-conditioned expression prediction.

Here we introduce DeepSpot-M (M for multimodal), a foundation model for predicting spatial gene expression from histology. It queries genes through biological embeddings derived from foundation models. Because genes are specified by embeddings rather than fixed output nodes, a single model can predict transcriptome-wide expression and generalise to genes not observed during training. We trained DeepSpot-M on a curated pan-cancer dataset comprising 730,000 paired histology-transcriptomic profiles across 14 cancer types and multiple institutions (Fig. 1A). Across held-out cancers, DeepSpot-M generalises zero-shot to unseen cancer types and outperforms specialised models trained directly on those cohorts. It predicts genes never used as training targets at accuracy comparable to observed genes. Through test-time adaptation, it extends to targeted single-cell spatial assays using as little as one slide. Applied to The Cancer Genome Atlas, DeepSpot-M generates a virtual spatial transcriptomics atlas across 32 cancer types, demonstrating deployment at population scale. Beyond standard expression prediction, the same query-based representation enables transcriptome restoration, cross-species non-coding RNA inference, in silico variant-effect mapping and natural-language querying without changing the model architecture (Fig. 1B,C). These capabilities together establish a scalable foundation for virtual spatial transcriptomics from routine histology.

**Fig. 1.**
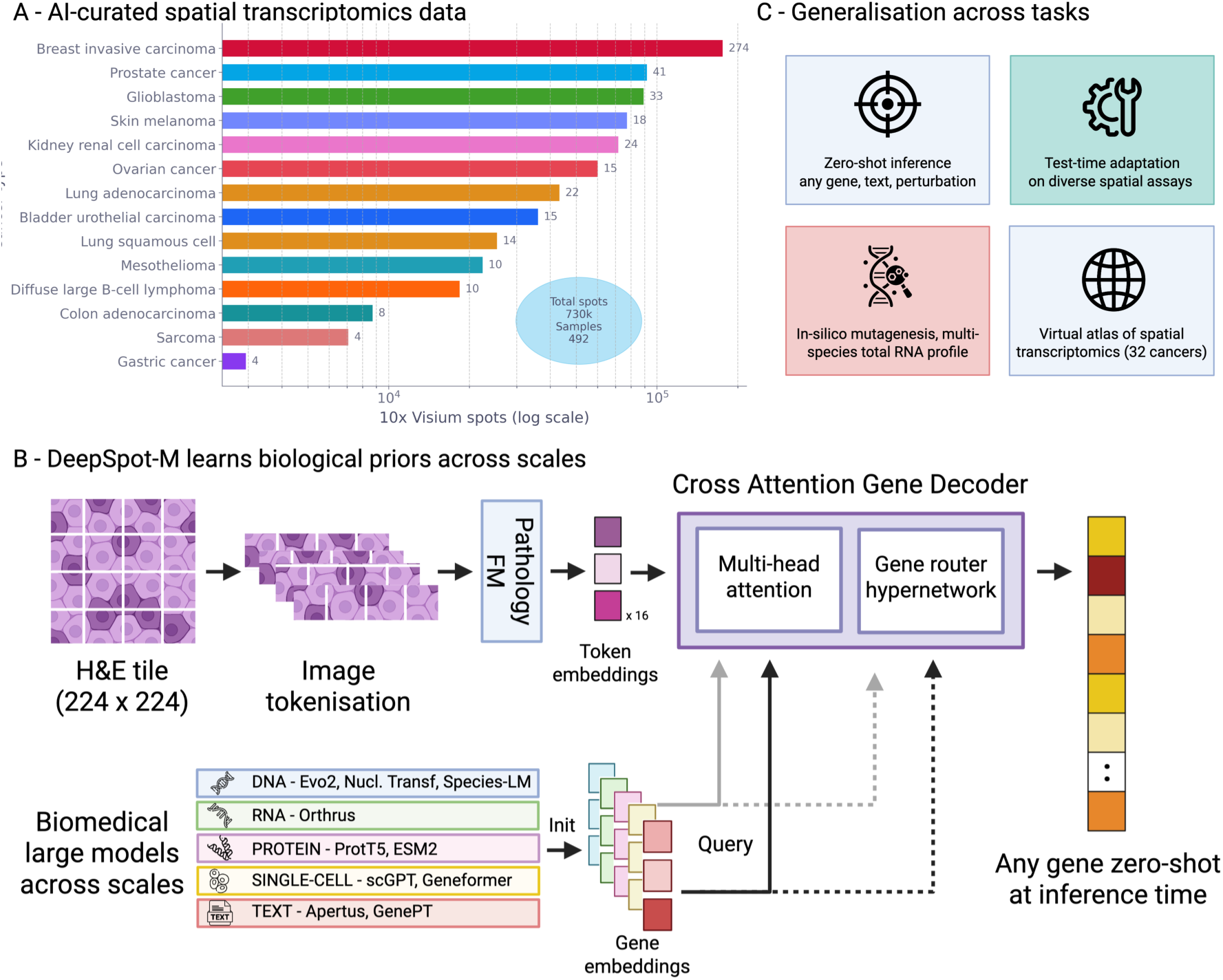
DeepSpot-M predicts transcriptome-wide spatial gene expression from histology. A) Training data from 15 10x Visium spatial transcriptomics datasets spanning 14 cancer types. B) DeepSpot-M architecture. A 224×224 H&E tile is tokenised into spatial patch embeddings by a LoRA-adapted pathology foundation model. A cross-attention gene decoder lets each gene query independently attend to patch tokens via multi-head attention, and a gene router hypernetwork generates gene-specific output projections from frozen biological embeddings drawn from DNA, RNA, protein, single-cell and text foundation models. This design enables zero-shot prediction of genes at inference time. C) Generalisation across tasks, including zero-shot gene inference and natural-language querying, one- and few-shot test-time adaptation to new spatial assays, in silico mutagenesis and spatial variant-effect mapping, and pan-cancer or cross-species inference using transfer learning.

## 2 Results

### 2.1 DeepSpot-M predicts transcriptome-wide spatial gene expression from histology

We first assembled a harmonised oncology spatial transcriptomics dataset pairing high-resolution H&E images with spot-level transcriptomes, comprising 730,000 paired histology-transcriptomic profiles from ∼500 10x Visium samples spanning 14 cancer types and multiple institutions (Fig. 1A, Extended Data Fig. 1). To our knowledge, this is among the largest and most diverse oncology ST datasets assembled to date. A unified preprocessing workflow (Section 4.2) harmonises the cohorts and is designed to reduce technical noise and batch effects. Its scale and diversity let DeepSpot-M learn morphology-expression relationships that generalise across cohorts rather than reflecting cohort-specific technical artefacts.

DeepSpot-M uses this dataset to learn a query-based mapping from local morphology to gene-specific expression (Fig. 1B). Instead of regressing a fixed panel of genes, the model predicts expression for each gene from a learned biological representation. Two design choices enable this formulation.

First, each gene is represented by embeddings from foundation models trained on complementary biological modalities, including DNA, RNA, proteins, single cells and biomedical text. These embeddings carry gene-level priors about sequence-function relationships, cellular states and gene-disease semantics. Because the underlying models are trained on large, diverse corpora outside the spatial domain, these priors are computed independently of the spatial assay and are therefore less affected by spot-level contamination and low capture efficiency.

Second, a cross-attention gene decoder lets each gene query attend directly to the spatial patch tokens from a pathology foundation model. Instead of summarising the tile as a single pooled embedding, such as the CLS token or the mean over patch tokens, each gene attends to the patches most relevant to it, producing a spatial gene-specific readout of local morphology. Because genes are represented as queryable embeddings rather than fixed outputs, DeepSpot-M predicts genes never seen during training zero-shot, without retraining or architectural changes. The same property also enables test-time adaptation, in which a sample’s own measurements supervise the model and let it recover what was not measured, adapting to a new assay from as little as one slide (Fig. 1C).

Together, this curated dataset and query-based architecture extend histology-based prediction from the few hundred genes accessible to prior methods to the entire protein-coding transcriptome, directly from routine histology.

### 2.2 DeepSpot-M outperforms specialised models across held-out cancer cohorts

With the aim of testing whether DeepSpot-M transfers to cancer cohorts not seen during training, we benchmarked it on five 10x Visium datasets from the multi-centre MOSAIC Window program [28], comprising 60 samples, 10-15 per cohort. Four cohorts represented cancer types absent from training, and one glioblastoma cohort tested transfer to an independent cohort of a cancer type present in the training data (Fig. 2A).

**Fig. 2.**
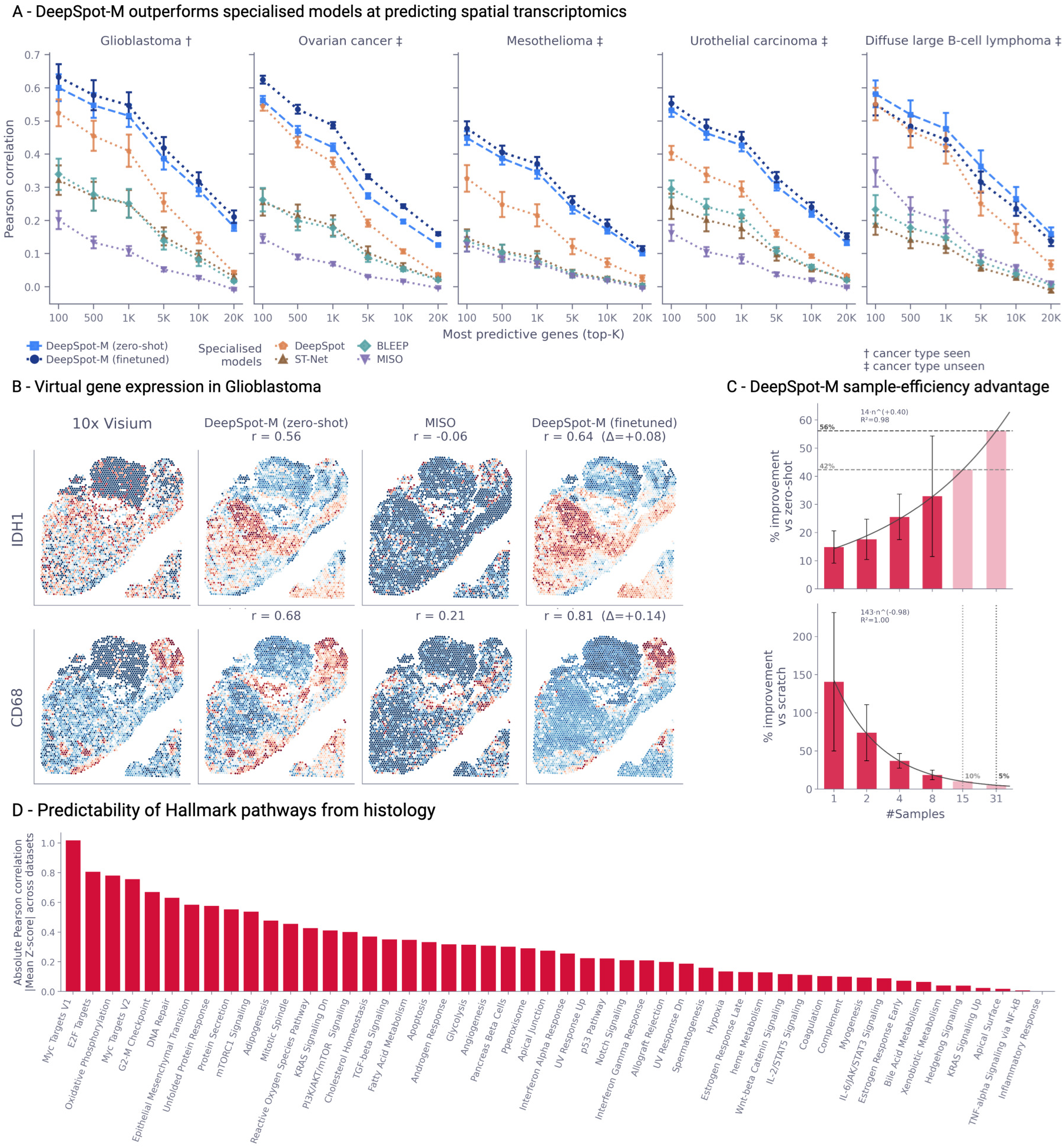
DeepSpot-M outperforms specialised models across held-out cancer cohorts. A) Benchmark of DeepSpot-M against specialised spatial methods on five MOSAIC Window 10x Visium cohorts, each labelled held-out (absent from training) or seen (present as a second, independent dataset). The evaluated cohort is always held out. Mean Pearson correlation between predicted and measured expression is shown against the number of top-predictive genes (N). B) Virtual spatial transcriptomics of a held-out glioblastoma sample for *IDH1* and *CD68*. C) Multi-cancer pretraining improves sample efficiency over zero-shot and scratch-trained baselines. Dark red bars are tested sample sizes, light red bars are extrapolated from the fitted scaling law. D) Zero-shot predictability of the 50 MSigDB Hallmark pathways [32] from H&E across 15 datasets, ranked by absolute mean Pearson correlation Z-score.

We compared DeepSpot-M with four specialised methods trained separately on each target cohort (ST-Net [13], BLEEP [16], MISO [15] and DeepSpot [14]). Performance was measured as gene-wise Pearson correlation between predicted and measured expression, as in prior work. We evaluated DeepSpot-M both zero-shot and after finetuning on the same cohort-specific data used by the baselines.

Zero-shot DeepSpot-M outperformed the specialised baselines across all cohorts, even though each baseline was trained directly on the cohort used for evaluation (Fig. 2A). On ovarian cancer, for example, DeepSpot-M improved the top-1,000-gene mean Pearson correlation by 14%, from 0.37±0.02 for the best baseline to 0.42 ± 0.01, with individual genes exceeding 0.60 within the top-100 set.

The advantage increased as more genes were evaluated. Specialised baselines declined sharply beyond the genes they predict best, whereas DeepSpot-M sustained positive correlations across thousands of additional genes. This widened the relative improvement to 43% for the top 5,000 genes and roughly twofold for the top 10,000 genes (Fig. 2A). By pretraining on a large, harmonised pan-cancer ST dataset, DeepSpot-M maintains predictive accuracy across a broader gene range than cohort-specific models trained on smaller datasets.

Finetuning further improved performance across gene sets (Fig. 2A). In ovarian cancer, the top-1,000-gene mean Pearson correlation increased to 0.51, a 22% improvement over zero-shot DeepSpot-M and a 38% improvement over the best baseline. Cross-cohort pretraining and cohort-specific finetuning are therefore complementary, with pretraining supporting zero-shot transfer and finetuning refining predictions for cohort-specific molecular programmes. For mesothelioma, the relative ranking of methods was preserved, though absolute performance was slightly lower, consistent with the smaller number of available spots and reduced image focus in this cohort (Extended Data Figs. 1 and 2). Across the benchmark, DeepSpot-M’s advantage over the specialised baselines persisted as more cohort-specific finetuning samples were added (Fig. 2C, Extended Data Fig. 3).

Spatial maps showed that the quantitative gains corresponded to interpretable predictions. In a held-out glioblastoma sample, zero-shot DeepSpot-M reproduced measured spatial gene expression of *IDH1*, a canonical glioma biomarker [29], and *CD68*, a marker of tumour-associated macrophages and activated microglia [30, 31] (Fig. 2B). Correlations were 0.56 and 0.68, respectively, whereas the specialised baseline (MISO [15]) failed to recover *IDH1* (−0.06). Finetuning increased these values to 0.64 and 0.81, sharpening the spatial signal so that the resulting maps more closely matched tumour and immune-infiltration patterns.

DeepSpot-M was also sample efficient, needing only a few finetuning slides to adapt to a new cohort (Fig. 2C). We assessed this by finetuning on increasing numbers of cohort-specific samples and tracking gains against two references, its own zero-shot predictions and a model trained from scratch on the same slides. Gains over the zero-shot model increased with finetuning sample size following a power law fit across the five MOSAIC Window cohorts (14 *n*^0.40^, *R*^2^ = 0.98), from 15% with one sample to an extrapolated 42% and 56% at 15 and 31 samples. Conversely, the advantage of pretraining over training from scratch decayed with sample size, again across the five cohorts (143 *n*^−0.98^, *R*^2^ = 1.00). One cohort-specific sample yielded a 140% improvement, with an extrapolated decrease to 10% at 15 samples and 5% at 31 samples. Pretraining helps most in the low-data regime, reducing the need for dozens of cohort-matched ST samples before accurate cohort-specific prediction becomes feasible.

Finally, we evaluated zero-shot predictability at the pathway level across the 50 MSigDB Hallmark gene sets [32] (Fig. 2D). Proliferation and cell-cycle programmes, including MYC, E2F, G2-M checkpoint and DNA repair, were most consistently recovered across cohorts. Oxidative phosphorylation, mTORC1 signalling and epithelial-mesenchymal transition were also highly predictable. By contrast, tissue-specific differentiation programmes and selected signalling pathways showed lower mean scores across cohorts, consistent with weaker or more context-dependent morphological associations in the zero-shot setting. These programmes may therefore require cohort- or tissue-specific finetuning to recover reliably.

### 2.3 DeepSpot-M predicts genes not observed during training

Having established cross-cohort transfer, we next asked whether DeepSpot-M can predict genes that were never used as training targets. This directly tests the query-based formulation, designed to let the model infer genes it never saw in training from their position in the shared molecular space (Fig. 1B).

To test this, we followed the chromosome-based holdout strategy established in prior work [33] and excluded all genes on chromosomes 1, 3, 5, 7 and 9, together with their paralogues, from the training targets to reduce information leakage. Afterwards, we evaluated these genes in independent patient cohorts. DeepSpot-M achieved comparable accuracy on excluded and observed genes, with overlapping per-gene correlation distributions (Fig. 3A), indicating that it learns transferable gene-level structure rather than memorising gene-specific mappings. Accuracy showed little dependence on a gene’s distance to its nearest training gene in embedding space, suggesting that DeepSpot-M exploits broader gene-gene structure rather than nearest-neighbour interpolation (Extended Data Fig. 4).

**Fig. 3.**
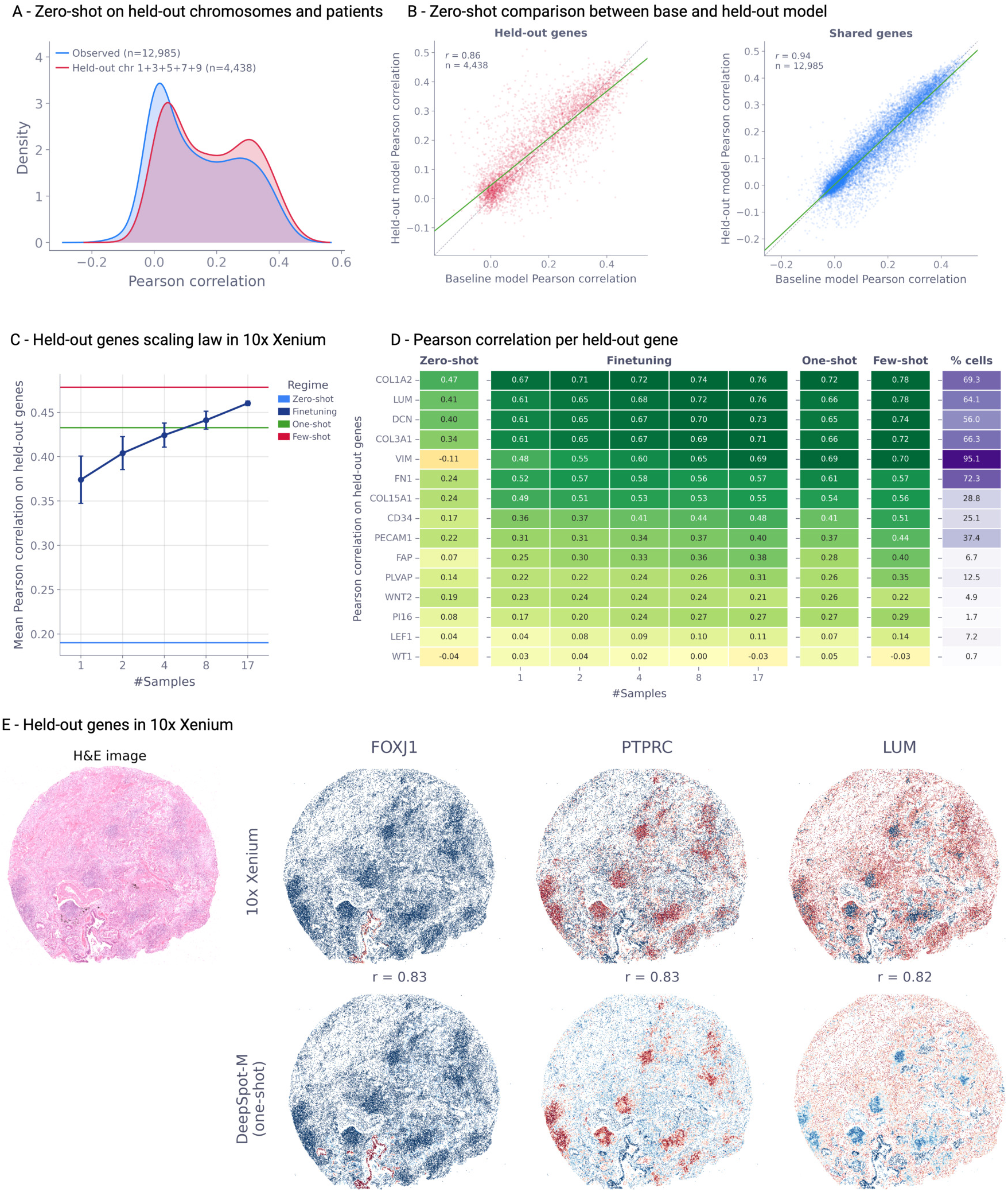
DeepSpot-M predicts genes unseen during training and extends targeted single-cell panels. A) Distribution of per-gene zero-shot Pearson correlations for observed genes (n = 12,985) and held-out genes from chromosomes 1, 3, 5, 7 and 9 (n = 4,438), evaluated on held-out patients from the MOSAIC Window program. B) Per-gene Pearson correlation of the gene-holdout model (y-axis) versus the full model trained on all genes (x-axis), shown for held-out genes (left) and shared genes (right). Each point is a gene. C) Scaling law on 10x Xenium. Mean Pearson correlation on held-out genes is shown versus the number of finetuning samples across zero-shot, finetuning, and one- and few-shot test-time adaptation regimes. D) Per-gene Pearson correlation for 15 held-out 10x Xenium genes. The rightmost column gives the percentage of cells expressing each marker in the Xenium reference. E) H&E image of a human lung 10x Xenium sample [35], followed by predicted spatial expression for three held-out genes from our virtual single-cell transcriptome-wide atlas alongside the matched 10x Xenium ground truth. These are *FOXJ1* (r = 0.83), a master regulator of motile ciliogenesis in ciliated airway epithelium, the pan-leukocyte marker *PTPRC* (CD45, r = 0.83) central to immune-cell signalling, and the fibroblast matrix proteoglycan *LUM* (lumican, r = 0.82).

In addition, we also compared the gene-holdout model with a full model trained on all genes. Their predictions agreed strongly for excluded genes (Pearson *r* = 0.86) and shared genes (Pearson *r* = 0.94), despite being trained with different gene vocabularies (Fig. 3B). The full model was more accurate for highly predictable genes, whereas the gene-holdout model performed better for less predictable genes (Extended Data Fig. 5). Notably, this pattern is consistent with the full model benefiting from direct gene-specific supervision, while the holdout model relies more strongly on gene-gene structure encoded in the biological embeddings. This reliance may be especially valuable for noisy or sparse measurements, where direct gene-specific supervision is less reliable.

DeepSpot-M can therefore accommodate heterogeneous gene vocabularies and predict genes absent from training, extending spatial expression inference beyond the genes measured in a given assay.

### 2.4 DeepSpot-M extends targeted panels at single-cell resolution

Next, we tested whether gene-level generalisation extends from spot-level 10x Visium (∼55 *µ*m) to single-cell-resolution 10x Xenium. Xenium measures a targeted panel that is fixed at acquisition, typically 300-500 genes and up to ∼5,000 in recent versions. This panel covers only a fraction of the transcriptome, and some genes are difficult to probe, such as those with sequence homology to related genes or high GC content [34]. A model that can fill in these unmeasured genes from the measured panel would substantially increase the molecular coverage of retrospective single-cell spatial data, extending targeted assays toward transcriptome-wide analysis without expanding the experimental panel.

More specifically, we evaluated DeepSpot-M on a public lung 10x Xenium dataset [35] of 315 genes across 20 patients, holding out three patients for testing. On these test patients, we predicted 15 held-out genes per cell from an H&E tile centred on its 10x Xenium nucleus centroid. We compared zero-shot prediction, finetuning on external Xenium samples, and one- and few-shot test-time adaptation that reuses the sample’s own measured panel as supervision (Fig. 3C).

Without single-cell training data, DeepSpot-M achieved a zero-shot mean Pearson correlation of 0.19 across the 15 genes. The stromal genes *COL1A2* and *LUM* reached 0.47 and 0.41, respectively (Fig. 3C,D). Finetuning on one external Xenium sample doubled mean held-out-gene accuracy to 0.38, with *COL1A2* reaching 0.67 and *LUM* reaching 0.61. Accuracy increased further with additional samples, and the most predictable genes reached correlations of 0.76. By contrast, poorly predicted genes such as *WT1* were largely absent from the test samples (expressed in 0.7% of cells), which limited both their recovery and their evaluation (Fig. 3D).

Test-time adaptation can reuse the genes a Xenium sample already measures as supervision, recovering the genes the panel leaves out without any external data. One-shot test-time adaptation on a single target sample more than doubled the zero-shot accuracy, reaching a mean Pearson correlation of 0.44 and matching finetuning on about eight external Xenium samples (Fig. 3C,D). Few-shot test-time adaptation, with a few additional same-assay samples, raised accuracy by a further 11%, surpassing a model finetuned on 17 external samples.

This efficiency is consistent with the organisation of gene expression into shared co-expression programmes and cell-type signatures, which DeepSpot-M already encodes through its biological gene embeddings, including those from single-cell foundation models. Test-time adaptation then anchors these priors to a sample’s own measured genes, inferring the unmeasured, co-regulated ones from the programmes that are active in that sample.

Consistent with this, across retrospective Xenium samples in HEST-1K [36], spanning 22 tissue types, per-tissue accuracy on observed and held-out genes was positively correlated (Pearson *r* = 0.94, *n* = 22 tissues, Extended Data Fig. 6). Tissues with higher measured-panel accuracy also tended to have higher held-out-gene accuracy. Spatial maps for three held-out genes spanning distinct cell compartments closely matched their ground-truth patterns (all *r* ≥ 0.82, Fig. 3E). These were the ciliated-epithelial marker *FOXJ1*, the pan-leukocyte marker *PTPRC* (CD45) and the fibroblast proteoglycan *LUM*.

DeepSpot-M adapts to a new spatial assay from as little as one slide and improves as target-specific data accumulate. By using a sample’s own panel as supervision, the model reduces dependence on large targeted panels and extends single-cell spatial assays toward transcriptome-wide coverage. We release virtual single-cell transcriptome-wide profiles for all 59 Xenium samples in HEST-1K, spanning 22 tissue types, as a public resource.

### 2.5 DeepSpot-M reconstructs bulk transcriptomes from histology

Beyond spatial ground truth, MOSAIC Window measures bulk RNA-seq for the same tissue [28], an independent assay averaged over the whole specimen. Reconstructing it from spatial predictions therefore tests whether DeepSpot-M generalises to an orthogonal molecular assay. We aggregated each sample’s predicted spatial expression into a pseudo-bulk profile and compared it with both the matched ST pseudo-bulk and bulk RNA-seq. Predictions aligned more closely with the ST pseudo-bulk than with bulk RNA-seq (top-100-gene Pearson correlation of 0.87 versus 0.82 in ovarian cancer, Extended Data Fig. 7). This is expected, because ST is directly paired to the input histology image, whereas bulk RNA-seq is a matched but coarser tissue-level measurement. The correlation between the two measured assays is itself imperfect and sets an approximate upper bound on how well bulk can be reconstructed (dashed grey line, Extended Data Fig. 7).

We also compared DeepSpot-M with Sequoia [37], a specialised histology-to-bulk model trained separately for each cancer type on TCGA. Although DeepSpot-M was never trained on bulk RNA-seq, in the zero-shot setting it matched or exceeded Sequoia across all five cohorts and approached this upper bound (ovarian cancer, 0.82 versus 0.80, Extended Data Fig. 7; Sequoia evaluation protocol in Supplementary Methods).

In essence, learning spatial gene expression captures molecular programmes that remain informative after aggregation to the tissue level. Consequently, a model trained solely on spatial transcriptomics can approximate bulk expression without bulk-specific supervision, while retaining the spatial information that bulk assays inherently discard.

### 2.6 DeepSpot-M generates a virtual spatial transcriptomics atlas across 32 cancers

Having established cross-cohort, cross-gene, cross-platform and bulk-level generalisation, we next tested whether DeepSpot-M can scale to population-level pathology archives. We applied it to The Cancer Genome Atlas (TCGA), generating a virtual spatial transcriptomics atlas of 28,664 whole-slide images (17,372 fresh-frozen and 11,292 formalin-fixed paraffin-embedded, FFPE), 295.3 million spots and 10,865 patients across 32 cancer types (Fig. 4A). To illustrate downstream use, we scored every spot for malignancy by applying CancerFinder [38], a domain-generalised malignant-cell classifier, to the predicted expression. We first validated these scores against expert malignant-versus-normal annotations across four independent datasets [14, 39, 40]. CancerFinder distinguished malignant from non-malignant tissue with high accuracy (AUROC 0.87-0.96 per dataset, 0.93 pooled across 87,998 annotated regions, Extended Data Fig. 9a), with malignancy scores significantly higher in malignant than normal regions in every dataset (Wilcoxon rank-sum test, *P <* 0.001, Extended Data Fig. 9b), confirming that the predicted scores reflect genuine tissue state rather than classifier artefacts. Visualising these scores across the atlas, a UMAP of the predicted profiles resolved malignant from non-malignant regions across cancers, recovering a pan-cancer map of malignancy directly from routine histology (Fig. 4B). The same embedding also organised spots by cancer type and by organ system, indicating that the predicted profiles retain cancer-type- and organ-specific structure (Extended Data Fig. 8). Notably, non-malignant spots tended to integrate into a shared compartment across organ systems, whereas malignant spots separated into organ- and cancer-type-specific regions.

**Fig. 4.**
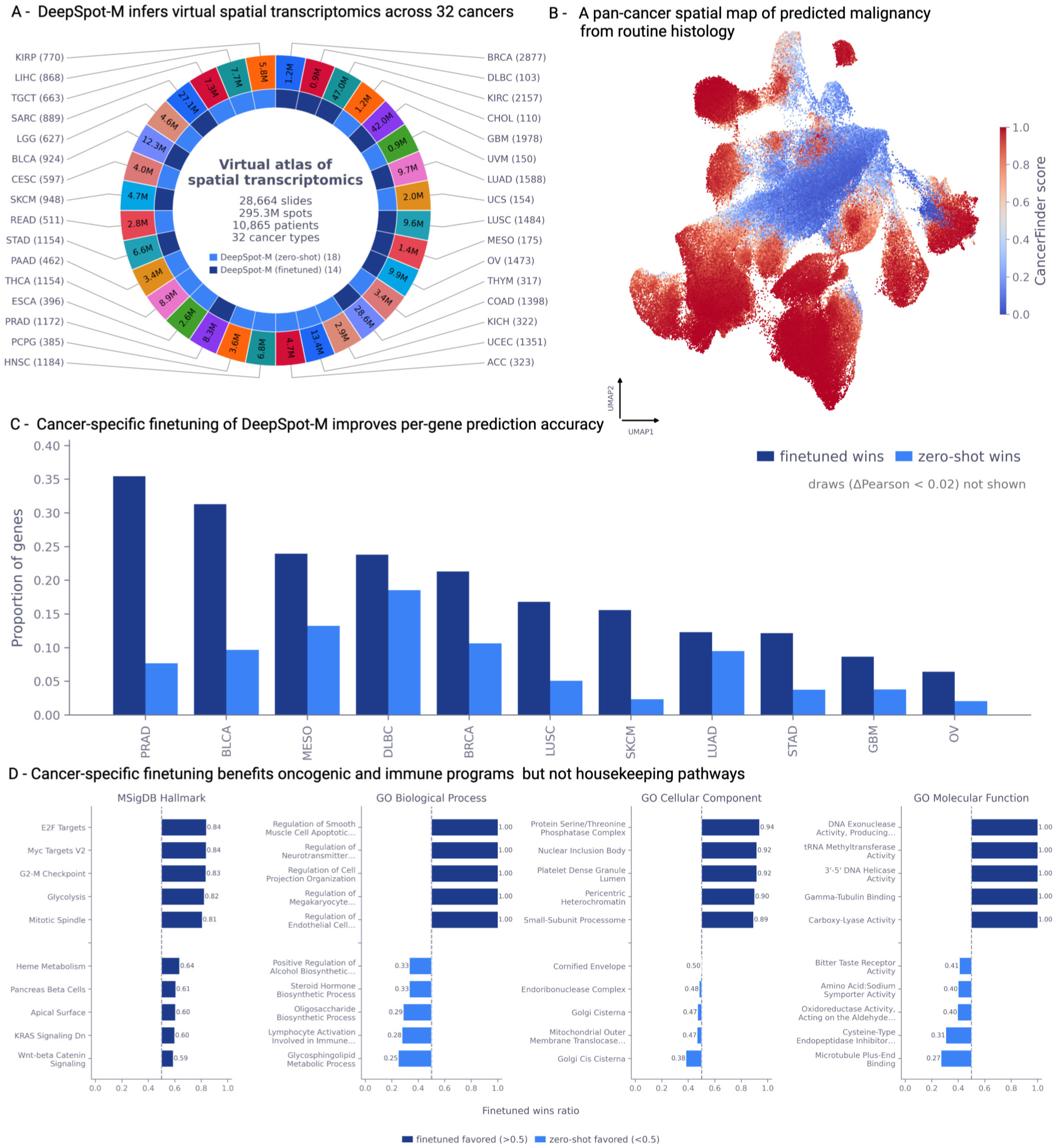
DeepSpot-M generates a virtual spatial transcriptomics atlas across 32 TCGA cancers. A) Virtual spatial transcriptomics atlas generated by applying DeepSpot-M to TCGA, comprising 28,664 slides (17,372 fresh-frozen and 11,292 FFPE), 295.3M spots and 10,865 patients across 32 cancer types. Predictions are zero-shot for 18 cancers and cancer-specific finetuned for 14 cancers. Values indicate the number of spots per cancer type. B) Pan-cancer spatial map of predicted malignancy from routine histology. Each point is a spot in a UMAP of predicted expression, coloured by predicted malignancy score. C) Cancer-specific finetuning improves per-gene accuracy. The plot shows the proportion of genes for which the finetuned (dark) or zero-shot (light) model achieves the higher Pearson correlation per cancer. Draws (ΔPearson < 0.02) are not shown. D) Finetuned versus zero-shot win ratio per pathway across MSigDB Hallmark and Gene Ontology gene sets. Ratios above 0.5 favour finetuning, and ratios below 0.5 favour zero-shot prediction.

Beyond the malignancy map, we assessed gene-level accuracy. Expression was predicted zero-shot for 18 cancer types without matching ST training data and with cancer-specific finetuning for the remaining 14. Although TCGA lacks spatial transcriptomic ground truth, matched bulk RNA-seq provides an orthogonal cohort-level check. For each slide, we aggregated predicted spatial expression into a pseudo-bulk profile and correlated it gene by gene with matched TCGA bulk RNA-seq across slides. Finetuning improved a larger fraction of genes in most cancers. In prostate cancer, for example, finetuned predictions outperformed zero-shot predictions for 32% of genes, compared with 16% favouring zero-shot prediction (Fig. 4C). Sarcoma was the main exception, likely because only four lower-quality ST samples were available for cancer-specific finetuning.

Stratifying these per-gene comparisons by pathway revealed where finetuning helps (Fig. 4D). Finetuning preferentially improved oncogenic and immune programmes, including reactive oxygen species, mTORC1 signalling, DNA repair and CD4 T-cell regulation. Conserved housekeeping and core metabolic programmes, such as mitochondrial electron transport and oxidative phosphorylation, were already well predicted zero-shot and gained little. Cancer-specific finetuning therefore sharpens predictions for disease-relevant, context-dependent programmes while leaving broadly conserved expression largely unchanged.

Beyond malignancy, we asked whether the atlas captures clinically relevant features of the tumour microenvironment at gene-level spatial resolution. We examined tertiary lymphoid structures (TLS), organised immune aggregates that mark active anti-tumour immunity and predict response to immunotherapy [41]. Across bladder, kidney, lung squamous and melanoma cohorts (BLCA, KIRC, LUSC and SKCM), predicted expression of canonical TLS and B-cell markers, including *CXCL13*, *MS4A1* and *CD79A*, was significantly higher within TLS than in non-TLS tissue (Wilcoxon rank-sum test, *P <* 0.001, Extended Data Fig. 10). The atlas therefore captures clinically relevant immune architecture from H&E at population scale.

Together, these results show that DeepSpot-M scales from individual spatial transcriptomics studies to population-scale pathology archives. Despite the absence of spatial transcriptomic ground truth, the resulting atlas agrees with expert malignancy annotations, reproduces matched bulk RNA-seq, captures cancer-specific molecular programmes and recovers clinically relevant immune structures such as tertiary lymphoid structures. Large histology cohorts can therefore be transformed into virtual spatial transcriptomic resources, enabling spatial molecular analyses across thousands of patients at a scale that remains impractical with experimental assays alone.

### 2.7 DeepSpot-M restores degraded spatial transcriptomes

The same query-based representation also supports applications where the measured transcriptome is incomplete or degraded, and where genes are specified by molecular or textual queries rather than by measurement. We first tested this in low-quality ST data, where measured expression can be too sparse or contaminated for downstream interpretation.

Spatial transcriptomics platforms such as 10x Visium are affected by low gene-detection sensitivity, gene dropout and spot-level contamination, which can distort downstream analyses [42–44]. These limitations are compounded in archival samples, where degraded RNA further reduces data quality.

We examined a kidney cancer sample [14] that had been excluded from analysis because key markers failed quality control (Fig. 5A). Regions annotated as tertiary lymphoid structures (TLS) lacked expression of established TLS markers [45]. Instead, marker expression appeared in spurious locations. Raw 10x Visium therefore identified TLS with an AUROC of only 0.53 (Fig. 5B).

**Fig. 5.**
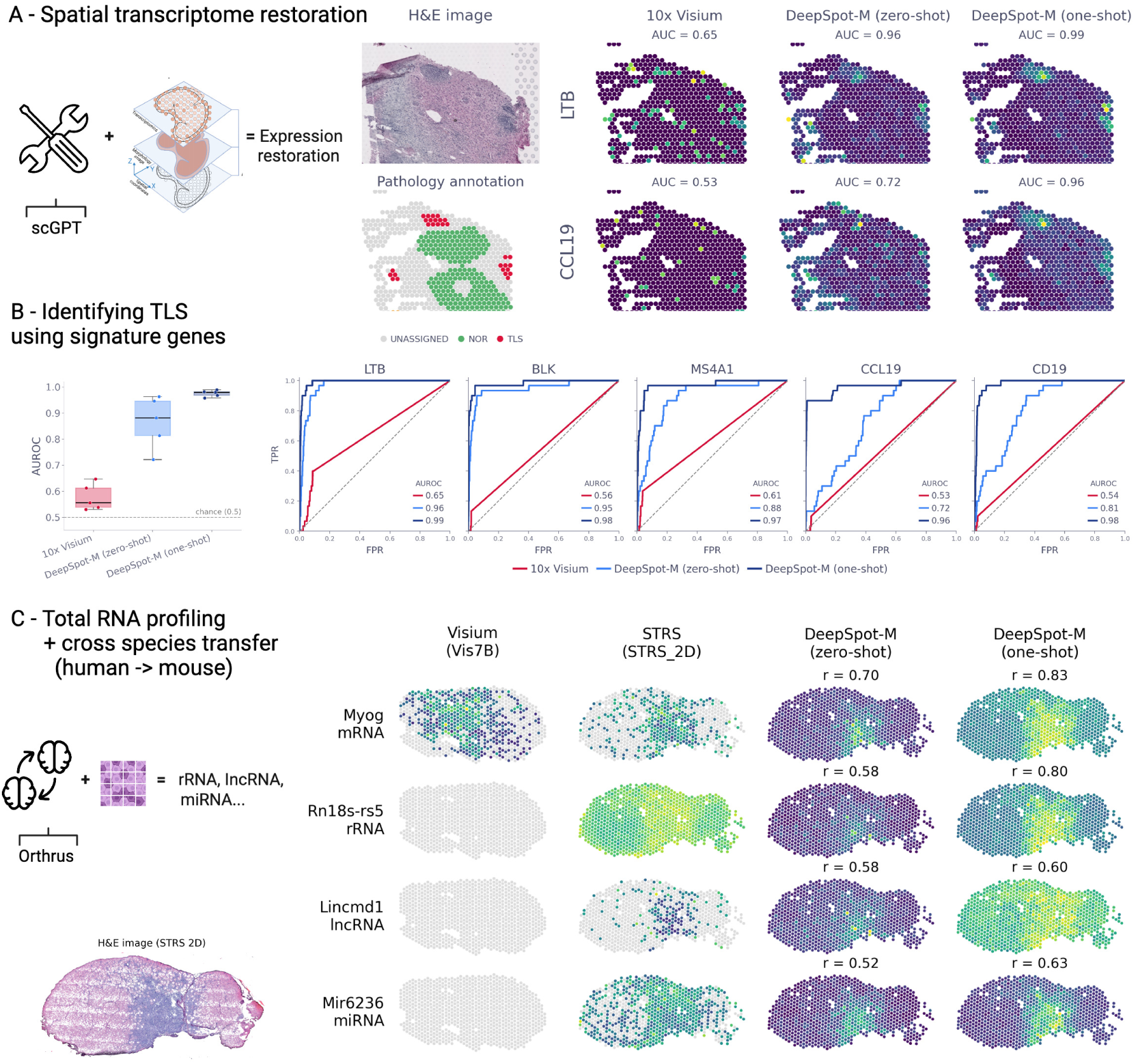
DeepSpot-M restores degraded spatial transcriptomes and profiles total RNA across species. A) Spatial transcriptome restoration in a low-quality 10x Visium kidney cancer sample. Spatial expression of the TLS markers *LTB* and *CCL19* is shown for raw 10x Visium, zero-shot DeepSpot-M and one-shot DeepSpot-M, together with the H&E image and pathology annotation. B) Identification of tertiary lymphoid structures from TLS signature genes. AUROC across samples (left) and per-gene ROC curves for *LTB*, *BLK*, *MS4A1*, *CCL19* and *CD19* compare 10x Visium, zero-shot DeepSpot-M and one-shot DeepSpot-M. B) Total RNA profiling and human-to-mouse transfer validated against STRS ground truth. Using RNA embeddings from Orthrus, DeepSpot-M infers coding and non-coding transcripts in mouse, including *Myog* (mRNA), *Rn18s-rs5* (rRNA), *Lincmd1* (lncRNA) and *Mir6236* (miRNA), in zero-shot and one-shot test-time adaptation settings, alongside the H&E image.

DeepSpot-M recovered TLS-marker expression zero-shot from the H&E image, raising the AUROC to 0.88 (Fig. 5A,B). One-shot test-time adaptation on the highly variable genes that passed quality control further aligned the predictions to the sample and produced coherent predicted spatial structure, increasing the AUROC to 0.97. Because no high-quality molecular ground truth was available for this sample, this analysis evaluates concordance with expert pathology annotation rather than direct molecular recovery.

Notably, DeepSpot-M can recover biologically plausible marker patterns in degraded or archival samples. This recovery draws on co-expression structure carried by the external gene embeddings, which is degraded in the measured data but retained in the priors. Conditioning on tissue morphology then repositions this latent structure within its spatial context. Because TLS marks active anti-tumour immunity and predicts response to immunotherapy [41], their recovery may make degraded samples informative for immune phenotyping.

### 2.8 DeepSpot-M extends spatial inference to total RNA profiles

We next asked whether DeepSpot-M could recover transcript classes that conventional spatial assays do not comprehensively capture. Most ST platforms capture polyadenylated RNA. Spatial total RNA-sequencing (STRS), based on enzymatic in situ polyadenylation, broadens transcript coverage but remains limited by accessibility, protocol complexity and lower signal-to-noise than conventional mRNA assays [46]. As a result, many spatial datasets provide only a partial view of regulatory and non-coding RNA programmes.

We applied DeepSpot-M to a mouse hindlimb injury sample profiled by both 10x Visium and STRS [46]. Querying the model with RNA embeddings from Orthrus [20] enabled prediction of coding and non-coding transcripts (Fig. 5C). Although trained only on human data, DeepSpot-M transferred across species and recovered the conserved muscle-regeneration marker *Myog* zero-shot, with a Pearson correlation of 0.70 against STRS.

DeepSpot-M also predicted spatial patterns for transcript classes incompletely captured by conventional 10x Visium assays. Representative rRNAs, lncRNAs and miRNAs showed spatial patterns concordant with STRS ground truth despite never appearing in training (Fig. 5C). For example, the model localised the miRNA *Mir6236* zero-shot with a Pearson correlation of 0.52.

STRS captures protein-coding and non-coding species together, so we applied one-shot test-time adaptation using only the coding genes measured in the sample, a subset of the transcripts that standard poly(A) assays such as 10x Visium capture, while holding out the transcripts used for evaluation. This supervision improved accuracy for held-out transcripts across RNA classes, raising the coding *Myog* from 0.70 to 0.83 and the rRNA *Rn18s-rs5* from 0.58 to 0.80, with a smaller gain for the lncRNA *Lincmd1*, from 0.58 to 0.60 (Fig. 5C). Observed coding transcripts can therefore anchor the recovery of broader transcriptomic programmes beyond the assay’s capture space. By drawing on functional RNA priors, DeepSpot-M extends spatial inference beyond poly(A)-restricted assays and transfers non-coding RNA prediction across species.

### 2.9 DeepSpot-M performs in silico mutagenesis in tissue context

A longstanding goal in genetics is to predict how genetic variation propagates from molecular perturbation to tissue phenotype. Because DeepSpot-M links protein representations, gene expression and histology in a shared framework, we asked whether it could generate tissue-conditioned hypotheses about how variant-altered protein representations may shift predicted expression (Fig. 6A).

**Fig. 6.**
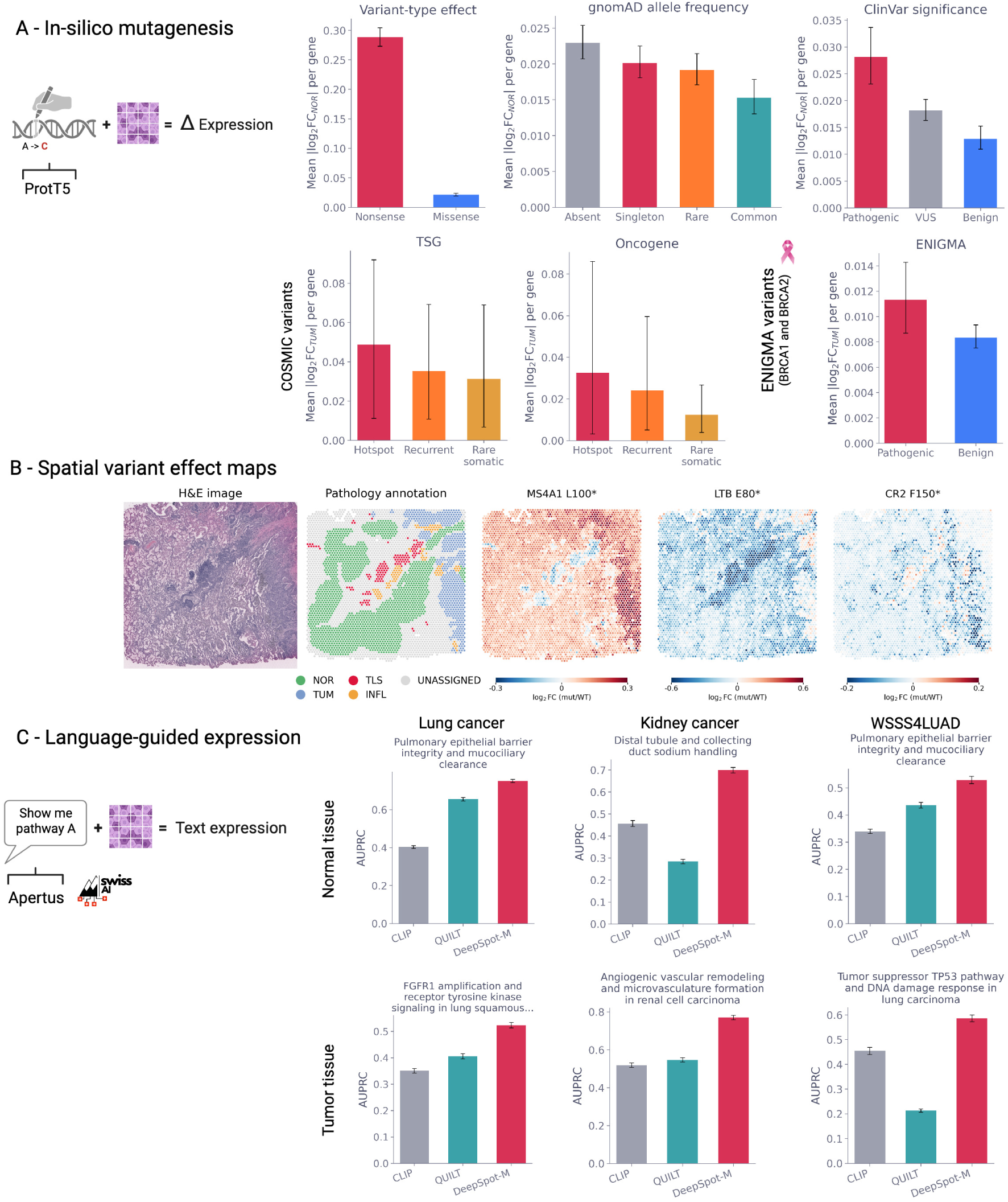
DeepSpot-M enables tissue-conditioned in silico mutagenesis and language-guided spatial transcriptomics. A) In silico mutagenesis conditioned on tissue architecture using protein representations from ProtT5. Mean | log_2_ FC| per gene is shown across variant annotations, including nonsense versus missense variants, gnomAD allele frequency, ClinVar significance, COSMIC variants in tumour suppressor genes (TSG) and oncogenes, and ENIGMA *BRCA1* /*BRCA2* variants. B) Spatial variant-effect maps. Predicted log_2_FC (mutant versus wild type) is shown across tissue patches for *LTB* E80*, *MS4A1* L100* and *CR2* F150*, together with the H&E image and pathology annotation. C) Language-guided spatial transcriptomics using text representations from Apertus. Area under the precision-recall curve (AUPRC) is shown for text queries in the USZ lung squamous cell carcinoma and clear cell renal cell carcinoma cohorts [14] and WSSS4LUAD, comparing CLIP, QUILT and DeepSpot-M in normal and tumour tissue.

For each variant, we regenerated the gene’s protein representation from its mutated amino-acid sequence (for example, a missense substitution or a nonsense truncation) and substituted it for the wild-type representation, then measured the resulting change in predicted spatial expression (log_2_ FC). Because predictions are conditioned on local morphology, this yields a tissue-conditioned estimate of a variant’s effect on expression, computed directly from H&E. We applied this procedure to both germline and somatic variants in a lung cancer sample (LC1 [14]). For the aggregate severity comparison (Fig. 6A), germline variants were evaluated in regions manually annotated as normal and somatic variants in regions manually annotated as tumour by an expert pathologist, consistent with their respective tissue origins, whereas the spatial variant-effect maps (Fig. 6B) were computed across all annotated regions.

For ranking variants by effect size, we summarised each variant by the magnitude of its predicted expression change (mean | log_2_ FC| between mutant and wild type), which captures effect size independently of the direction of the change. Across genes, this magnitude tracked established measures of variant deleteriousness along several independent axes. By variant type, nonsense variants produced substantially larger changes than missense variants, mirroring their expected severity (Fig. 6A). Population frequency showed the same trend, as the rarest variants in gnomAD [47], those absent or seen only once, had the largest predicted effects and common variants the smallest. Clinical classification agreed, with ClinVar [48] pathogenic variants exceeding variants of uncertain significance, which in turn exceeded benign variants.

This concordance extended from germline to somatic and clinically curated variation. In COSMIC [49], recurrent and hotspot somatic mutations in both oncogenes and tumour suppressor genes produced larger changes than rare somatic variants, and among ENIGMA-classified *BRCA1* and *BRCA2* variants [50], pathogenic variants once more exceeded benign ones (Fig. 6A).

To examine directionality, we then mapped the signed predicted expression change (log_2_ FC) for individual variants across tissue (Fig. 6B). Because each prediction reflects the local tissue context, predicted effects varied across the slide rather than being fixed per gene. The co-expressed B-cell markers *MS4A1* (CD20) and *CR2* (CD21) illustrate this context dependence. Truncating *MS4A1* (L100*) was predicted to increase expression in tumour regions but decrease it in TLS, whereas truncating *CR2* (F150*) showed the opposite pattern. These signed shifts describe predicted changes in spatial gene expression in tissue context and are not direct readouts of protein function. A variant may raise the predicted expression of a gene while remaining biologically deleterious, so the sign of the predicted change should not be equated with gain or loss of function.

These analyses suggest that variant-altered protein representations induce prediction shifts that align with variant-severity annotations and vary with tissue context. The resulting spatial variant-effect maps provide tissue-conditioned hypotheses without variant-specific training.

### 2.10 DeepSpot-M maps spatial expression from natural language

Natural language provides a flexible interface for describing biological processes. Linking free-text descriptions to spatial molecular maps, however, requires a shared representation across language, molecular programmes and histology.

We tested whether DeepSpot-M can map natural-language queries to spatial molecular maps across cancer types and tissue compartments (Fig. 6C). In a lung squamous cell carcinoma cohort from the University Hospital Zurich (USZ) [14], the query “pulmonary epithelial barrier integrity and mucociliary clearance” localised more strongly to normal than tumour tissue, whereas “FGFR1 amplification and receptor tyrosine kinase signaling in lung squamous cell carcinoma” showed the opposite. In a clear cell renal cell carcinoma cohort from USZ [14], “angiogenic vascular remodelling and microvascularisation in renal-cell carcinoma” localised to tumour regions. We observed the same pattern on the WSSS4LUAD dataset [40].

Across queries and cancer types, DeepSpot-M outperformed CLIP [51] and the pathology-specific language model QUILT [52]. For angiogenic vascular remodelling in renal-cell carcinoma, for example, DeepSpot-M achieved an AUPRC of 0.77 ± 0.01, compared with 0.52 ± 0.01 for CLIP and 0.55 ± 0.01 for QUILT (Fig. 6C). The model maps free-text biological concepts to spatial molecular maps aligned with tissue compartments, while remaining sensitive to query phrasing (CLIP and QUILT evaluation in Supplementary Methods).

## 3 Discussion

We introduced DeepSpot-M, a multimodal foundation model for predicting spatial gene expression from routine histology. The central design choice is to represent genes as biological queries rather than as fixed output nodes. This changes the prediction problem from fitting a closed gene panel to learning a morphology-conditioned readout over a shared molecular space. As a result, one model can span the protein-coding transcriptome, predict genes excluded from training and adapt to assays with different measured panels.

Trained on a large harmonised oncology ST dataset, DeepSpot-M generalised across cohorts, cancer types and platforms. In held-out cancers, it outperformed specialised models trained directly on the target cohorts, and its advantage was largest when paired ST data were scarce. This low-data regime is common for rare diseases, emerging spatial assays and new clinical cohorts. Test-time adaptation further showed that the measured genes of a single slide can anchor the recovery of unmeasured, co-regulated genes, extending targeted panels without requiring a dedicated cohort-scale training set.

DeepSpot-M differs from existing histology-to-expression methods in both how genes are represented and how the model is deployed. Supervised models that regress a fixed gene panel from a single cohort, including ST-Net [13], DeepSpot [14] and MISO [15], are bounded by the genes and the cohort seen in training. Retrieval-based methods such as BLEEP [16] and visual-omics models such as OmiCLIP [53] instead impute expression by matching each tile to its nearest neighbours in a reference of measured profiles. This requires access to that reference at inference, bounds predictions to profiles it already contains, and can be impractical for sensitive clinical data that cannot be shared alongside the model. Histology-to-bulk models such as Sequoia [37] recover cohort-level expression but discard spatial structure. By representing each gene as a parametric biological query rather than a fixed output or a reference lookup, DeepSpot-M predicts expression generatively from morphology alone, without a reference set at inference. This open, query-based formulation, rather than a larger fixed panel or reference, is the main difference from prior work.

The query-based representation also explains why the same architecture supports tasks beyond standard expression prediction. Foundation models trained on DNA, RNA, proteins, single cells and biomedical text encode complementary gene-level priors, including sequence-function relationships, cellular states, protein properties and gene-disease semantics. By conditioning these priors on local morphology, DeepSpot-M recovered marker patterns in degraded measurements, transferred to non-coding RNA across species, generated tissue-conditioned variant-effect hypotheses and mapped natural-language descriptions to spatial molecular maps. These tasks differ in the form of the query but share the same underlying mechanism, in which a biological representation queries which molecular programme is visible in a given histological context.

Predictive accuracy was not uniform across genes and pathways. Proliferation and cell-cycle programmes were most consistently recovered, whereas tissue-specific differentiation and selected signalling programmes were more context dependent. Cancer-specific finetuning preferentially improved oncogenic and immune programmes while leaving conserved housekeeping expression largely unchanged. These patterns suggest that DeepSpot-M captures the transcriptomic programmes most clearly reflected in tissue structure, and that finetuning refines the disease-specific component of this signal.

Several limitations define the current scope. Zero-shot prediction provides a strong starting point where no paired ST data exist. Where even a few matched slides are available, cohort-specific finetuning and test-time adaptation markedly improve accuracy, refining zero-shot maps into cohort-accurate predictions, and should be the preferred mode. Most training data are spot-level 10x Visium measurements, so richer single-cell spatial data should improve transfer to cellular resolution. Genes with weak spatial structure or little morphological correlate are inherently harder to infer from H&E, reflecting limited observable signal rather than model capacity alone. Reported accuracy also depends on the number of spots in a sample. Because each gene’s Pearson correlation is computed across these spots, fewer spots make a high correlation easier to reach, so smaller tissues tend to score higher and absolute values are not directly comparable across tissues of different size. These correlations are computed against a measured reference smoothed over each spot’s nearest neighbours in expression space, so absolute values depend on this choice. Across alternative smoothing settings and raw measured counts, the ranking of methods is unchanged (Extended Data Figs. 17, 18). Variant-effect maps should be treated as tissue-conditioned hypotheses, because the perturbation is introduced by replacing a wild-type protein embedding with a variant embedding and has not been validated in tissues carrying the corresponding mutation. Since the model predicts spatial gene expression rather than protein activity, the direction of a predicted expression change should not be equated with gain or loss of protein function. Language-guided maps likewise depend on the text representations and biological descriptions available to the model. Both capabilities require experimental or clinical validation before use in decision-making. Cross-species transfer should likewise be interpreted with care. *Myog*, a highly conserved muscle marker (95% human-mouse protein identity [54]) recovered in a muscle-rich hindlimb sample, is a favourable case, so transfer may be weaker for more divergent genes or sparsely represented tissues. Finally, although DeepSpot-M generalises across cancers, broader training data will be needed for markedly different non-malignant tissues and disease settings.

These limitations point to natural extensions. Paired multimodal datasets combining ST with whole-genome sequencing, pathology reports, protein readouts and richer single-cell assays could allow future models to relate genotype, histology, text and proteome in spatial context. Because DeepSpot-M queries genes through external biological embeddings, advances in the underlying foundation models should in turn sharpen prediction for the genes and programmes that remain hardest to infer, without changes to the architecture.

In the future, DeepSpot-M could be systematically applied to infer virtual spatial transcriptomics across large pathology archives, prioritise biomarkers in retrospective clinical cohorts and generate tissue-conditioned hypotheses linking genetic variation to spatial expression. To foster these applications, we release the trained model together with the TCGA virtual spatial transcriptomics atlas and virtual single-cell profiles for HEST-1K as open resources for biomarker discovery and for benchmarking virtual spatial transcriptomics models. We hope that DeepSpot-M will stimulate a deeper understanding of how molecular programmes shape tissue structure and facilitate the development of improved spatial biomarkers for precision oncology.

## 4 Methods

### 4.1 Spatial transcriptomics datasets

#### Pan-cancer corpus

We assembled a harmonised pan-cancer corpus of 15 10x Visium spatial transcriptomics datasets spanning 14 cancer types and multiple institutions, comprising ∼500 samples and 730,000 spot-level histology-transcriptomic profiles. Each profile pairs a high-resolution haematoxylin and eosin (H&E) tile with the quantitative transcriptome of the corresponding Visium spot (∼55 *µ*m). Whole-slide images were scanned at an average magnification of 20× (mean 0.50 *µ*m per pixel, ranging from ∼10× to ∼40× across datasets). Per-dataset spot counts, gene coverage, sequencing depth and scanning magnification are summarised in Extended Data Fig. 1.

#### Benchmark cohorts

The main cross-cohort benchmark uses five 10x Visium cohorts from the multi-centre MOSAIC Window program [28] (60 samples, 10-15 per cohort), comprising ovarian cancer, mesothelioma, diffuse large B-cell lymphoma, urothelial carcinoma and glioblastoma. These cohorts are included in the overall 15-dataset corpus, but each target cohort was removed completely from training in the corresponding leave-one-dataset-out experiment (see “Leave-one-dataset-out benchmarking”). Four benchmark cohorts therefore test transfer to cancer types absent from the training split, whereas the glioblastoma cohort tests transfer to an independent cohort of a cancer type represented by another dataset. Each cohort additionally provides matched bulk RNA-seq for the same specimen, used for the orthogonal pseudo-bulk benchmark.

#### Single-cell and additional datasets

Beyond the pan-cancer Visium corpus, DeepSpot-M was evaluated on a public lung 10x Xenium dataset [35] and the 59 HEST-1K 10x Xenium samples [36] for transfer to single-cell resolution, a mouse hindlimb sample profiled by both 10x Visium and spatial total RNA-sequencing (STRS) [46] for total-RNA and cross-species inference, a low-quality kidney cancer Visium sample [14] for transcriptome restoration, and the USZ lung squamous cell carcinoma and clear cell renal cell carcinoma cohorts [14] with WSSS4LUAD [40] for language-guided querying. Population-scale deployment used TCGA fresh-frozen and FFPE diagnostic whole-slide images. Sample sizes and splits for each dataset are given in the corresponding subsections.

### 4.2 Preprocessing and gene vocabulary

All cohorts passed through a unified preprocessing workflow that harmonises tiling, expression normalisation and quality control across platforms and institutions (Extended Data Fig. 11). H&E tiles were extracted at a consistent physical resolution and matched to spot coordinates. Spot-level expression was normalised to counts per 10,000 and log(1 +*x*)-transformed. Expression targets were aligned to a fixed vocabulary of 19,338 protein-coding genes from the Ensembl release 116 GRCh38 (hg38) annotation, and a binary gene mask recorded which genes were measured in each sample so that the loss and all metrics were computed only on measured genes. To reduce dropout noise, training targets combined raw measurements with spatially denoised expression (self-distillation from DeepSpot [14]), which improved accuracy across held-out cancers (Extended Data Fig. 11).

### 4.3 Biological gene embeddings

Each gene is represented by precomputed embeddings drawn from foundation models spanning five molecular modalities (DNA [17], RNA [20], protein [21], single cells [23] and biomedical text [27]). For each modality we benchmarked alternative pretrained sources as ablations, including DNA [18, 19], protein [22], single-cell [24] and text [26] models, against a random-embedding lower bound (Extended Data Fig. 12). All embeddings were pre-aligned to the gene vocabulary. Genes missing from a given modality were assigned that modality’s mean embedding with small additive noise to preserve uniqueness. Because these embeddings are computed independently of the spatial assay, they provide priors that are less sensitive to spot-level contamination and capture efficiency.

### 4.4 DeepSpot-M architecture

DeepSpot-M maps a single H&E tile to gene expression through a query-based decoder, so that genes are specified by biological embeddings rather than fixed output nodes. For tile *x_i_*, the model predicts an expression vector ŷ*_i_* ∈ R*^G^* over the active gene vocabulary. The architecture has three components.

#### Vision encoder

Tiles are tokenised into spatial patch embeddings by a pathology foundation model (Midnight [55]) kept frozen and adapted to the task with low-rank adapters (LoRA) on the attention projections, adding less than 0.5% trainable parameters (Extended Data Figs. 13, 14). The encoder produces patch tokens

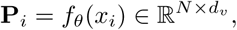

excluding the class token. A learned linear projection maps these tokens into the decoder dimension, **K***_i_* = **P***_i_***W***_p_* ∈ R*^N^*^×^*^d^*, which is used as both key and value in cross-attention.

#### Cross-attention gene decoder

Rather than pooling the tile into a single embedding, a cross-attention decoder lets each gene query attend directly to the patch tokens through stacked multi-head cross-attention blocks, producing a gene-specific spatial readout of local morphology (Extended Data Fig. 15). For gene *g* and biological source *m*, let 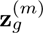 denote its precomputed biological embedding. A source-specific adapter *a_m_*(·) projects this embedding into the decoder space and, in multimodal models, adds a fixed modality code **e***_m_*:

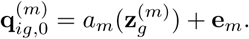

For layer *ℓ* = 1*, …, L*, each gene query is updated by cross-attention to the image patch tokens followed by a feed-forward block,

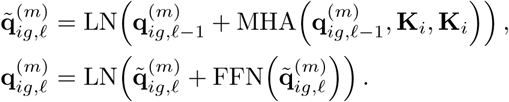

Thus, every gene obtains its own morphology-conditioned representation 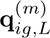.

#### Gene router hypernetwork

The output projection is generated per gene by a router hypernetwork that maps each gene’s biological embedding to its output weights, replacing a fixed per-gene head. For each active source *m*, two small multilayer perceptrons generate a gene-specific projection vector and bias,

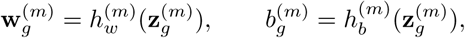

and expression is predicted as

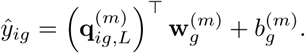

Because the router is conditioned on biological embeddings rather than on fixed output indices, it produces predictions for genes never seen in training, provided an embedding is available, and it improves accuracy on observed genes (Extended Data Fig. 16).

#### Multimodal training

At training time, one modality is sampled per batch in a deterministic round-robin and only its modality-specific adapter and router are updated, while the shared encoder and cross-attention decoder are updated on every batch. This exposes the backbone to every modality each epoch while keeping the per-batch cost equal to a single-modality model. At inference, any modality, or an ensemble across modalities, can query the decoder depending on which representation is available for the gene of interest.

### 4.5 Training

DeepSpot-M was trained to minimise a masked mean squared error on measured genes using the AdamW optimiser with a cosine-annealed learning-rate schedule and mixed precision. For measured expression *y_ig_*, prediction *y̅_ig_* and binary gene-availability mask *M_ig_*, the objective was

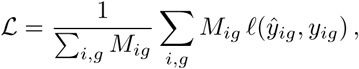

where *ℓ*(*y̅_ig_, y_ig_*) = (*y̅_ig_* − *y_ig_*)^2^ is the squared error. Per-source modality adapters, gene embeddings and routers were excluded from weight decay to preserve the biological prior under round-robin sampling. Data augmentation comprised random quality degradation (to emulate variable scan and capture quality), 90^◦^ rotations, horizontal and vertical flips, and colour jitter.

### 4.6 Evaluation metric and ground-truth correction

Predictive accuracy was reported as the gene-wise Pearson correlation between predicted and measured expression. For gene *g*, let Ω*_g_* denote the set of evaluation observations in which *g* was measured, and let *y̅_ig_* and *y_ig_* denote predicted and measured expression for observation *i*. We computed

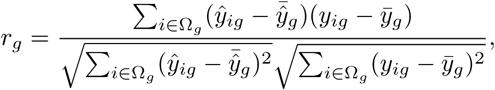

where 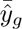 and *y̅_g_* are means over Ω*_g_*. Genes were ranked by *r_g_* within each dataset and comparison, and top-*K* performance was reported as the cumulative mean 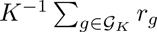, where *G_K_* contains the *K* highest-ranked genes.

Because 10x Visium measurements are affected by gene dropout, spot-level contamination and coarse spot resolution [43, 44], all reported gene-wise Pearson correlations were computed against a smoothed measured reference, averaging each spot’s measured expression over its *k* = 5 nearest neighbours in a PCA latent space of log-normalised ST expression. We also report correlations across additional neighbourhood sizes (*k* = 10, 15), Leiden clusters (resolutions 1, 0.5, 0.3, 0.2 and 0.1) and raw measured counts (Extended Data Figs. 17, 18).

### 4.7 Leave-one-dataset-out benchmarking

Cross-cohort generalisation was evaluated by leave-one-dataset-out (LODO). For each of the 15 datasets in turn, DeepSpot-M was trained on the remaining datasets and evaluated zero-shot on the held-out dataset, which was not used for model fitting in that split. The four benchmark cohorts with a unique cancer type therefore test transfer to unseen cancer types, whereas the glioblastoma cohort tests transfer to an independent cohort of a type present elsewhere in the training split. We compared against four specialised methods trained directly on each target cohort (ST-Net [13], BLEEP [16], MISO [15] and DeepSpot [14]). Pathway-level predictability was assessed across the 50 MSigDB Hallmark gene sets [32] by taking the mean per-gene correlation within each pathway and converting it to a Z-score for cross-dataset comparison.

### 4.8 Finetuning and sample-efficiency scaling

For adaptation to new cancer types or platforms, we resumed from a pretrained LODO checkpoint and finetuned the decoder together with the vision-encoder adapters, keeping the biological gene embeddings frozen so that small finetuning sets do not overwrite the prior. To quantify sample efficiency, we titrated the number of finetuning slides on the five MOSAIC Window cohorts and on the Xenium lung data, repeating each configuration across random seeds, and for each configuration compared the pretrained initialisation with a model trained from scratch on the same slides. Improvements as a function of finetuning sample size were summarised with power-law fits.

### 4.9 Test-time adaptation

Test-time adaptation reuses a sample’s own measured genes as supervision to recover the genes it does not measure. Starting from a pretrained checkpoint, the decoder is adapted on the measured panel of the target sample (one-shot) or of a few additional same-assay samples (few-shot). The biological embeddings remain frozen, so the unmeasured, co-regulated genes are inferred from the programmes active in that sample. This is the mechanism used for single-cell panel extension, transcriptome restoration and total-RNA inference.

### 4.10 Gene-holdout (chromosome) evaluation

To test prediction of genes never used as training targets, we adopted the chromosome-based holdout strategy established in prior work [33] and excluded all genes on chromosomes 1, 3, 5, 7 and 9, together with their paralogues on other chromosomes, from the training targets to reduce information leakage. Evaluation then compared *n* = 4,438 held-out genes on the excluded chromosomes with 12,985 observed genes used as training targets; the withheld paralogues were neither trained on nor evaluated as held-out genes, which accounts for the remaining genes in the 19,338-gene vocabulary. The loss was restricted to observed genes via the gene mask. Per-gene Pearson correlations were reported separately for held-out and observed genes on independent patient cohorts. We compared this gene-holdout model with a full model trained on all genes, and tested whether held-out accuracy was explained by a gene’s distance to its nearest training gene in embedding space (Extended Data Figs. 4, 5).

### 4.11 Transfer to single-cell resolution

Cross-platform transfer from Visium (∼55 *µ*m) to single-cell 10x Xenium was evaluated on lung data [35], holding out three patients and 15 panel genes for testing and comparing zero-shot prediction, finetuning on external Xenium samples, and one- and few-shot test-time adaptation that reuses the sample’s own panel. Single-cell predictions were generated by extracting an H&E tile centred on each nucleus centroid from the 10x Xenium segmentation and passing it through the model, and predicted expression was compared with the measured panel counts for that cell. Cross-tissue generalisation was assessed across the 59 HEST-1K Xenium samples, relating per-tissue accuracy on measured panels to accuracy on held-out genes. We release virtual single-cell transcriptome-wide profiles for all HEST-1K Xenium samples as a public resource.

### 4.12 Bulk transcriptome reconstruction

In order to test generalisation to an orthogonal assay, each sample’s predicted spatial expression was aggregated into a pseudo-bulk profile by averaging predicted expression over all in-tissue spots, 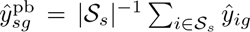, and correlated gene by gene with matched bulk RNA-seq across samples from the same cohort. The matched ST pseudo-bulk was computed analogously from the measured, normalised ST counts. We compared predictions with both the matched ST pseudo-bulk and bulk RNA-seq across the same top-*K* gene sets used in the spatial-expression benchmark, and benchmarked against pretrained Sequoia models [37], specialised histology-to-bulk models trained per cancer type on TCGA. The correlation between the two measured assays (ST pseudo-bulk versus bulk RNA-seq) provides a practical upper bound on reconstruction. Pseudo-bulk and bulk comparisons were computed at the sample level, separately from the within-sample spot-level correction schemes used for the spatial benchmark (see “Evaluation metric and ground-truth correction”, Extended Data Fig. 17).

### 4.13 TCGA virtual atlas

We applied DeepSpot-M to TCGA whole-slide images, comprising 17,372 fresh-frozen and 11,292 FFPE diagnostic slides (28,664 slides, 10,865 patients), which have no matched spatial transcriptomic data, so expression was predicted from the H&E image alone. For the 14 cancer types also represented in our spatial transcriptomics corpus, we used a model finetuned on the matching cancer-type ST cohorts. The remaining 18 cancer types have no corresponding ST data and were predicted zero-shot. Tissue tiles were separated from background before prediction. To illustrate downstream use, each spot was scored for malignancy by applying CancerFinder [38], a domain-generalised malignant-cell classifier, to the predicted expression, and predicted profiles were visualised with UMAP. Cohort-level validity was checked by correlating per-slide pseudo-bulk predictions with matched TCGA bulk RNA-seq, and gene-level wins of finetuned versus zero-shot prediction were stratified by MSigDB Hallmark [32] and Gene Ontology [56] gene sets. In order to probe tumour-microenvironment structure, predicted expression of canonical tertiary lymphoid structure and B-cell markers was compared between annotated TLS and normal regions (Wilcoxon rank-sum test, Extended Data Fig. 10), using the TCGA TLS annotations from [57].

### 4.14 Transcriptome restoration

For the degraded kidney cancer sample, DeepSpot-M predicted TLS-marker expression zero-shot from the H&E image, followed by one-shot test-time adaptation on the highly variable genes that passed quality control. Recovery of tertiary lymphoid structures was quantified as the AUROC of TLS-signature-gene expression against expert pathology annotation, comparing raw Visium, zero-shot and one-shot predictions.

### 4.15 Total RNA and cross-species inference

To recover transcript classes beyond poly(A)-selected mRNA, we queried the model with RNA embeddings from Orthrus [20] and predicted coding and non-coding transcripts (mRNA, rRNA, lncRNA and miRNA) in the mouse hindlimb sample, validating against STRS ground truth [46]. Mouse transcript sequences were obtained from Ensembl release 116 for *Mus musculus* (GRCm39; https://ftp.ensembl.org/pub/release-116/fasta/mus_musculus/). Although trained only on human data, the model transferred across species. One-shot test-time adaptation used only the coding genes measured in the sample, a subset of the transcripts that standard poly(A) assays capture, to anchor recovery of the broader transcriptome. Transcripts used for evaluation, including *Myog*, were excluded from the adaptation supervision. Human-to-mouse protein-sequence identity (95% for *Myog*) was computed by BLAST alignment of the orthologous protein sequences [54].

### 4.16 In silico mutagenesis

Variant effects were estimated by replacing a gene’s stored protein representation, which is its wild-type ProtT5 embedding [21], with the representation of its variant sequence. Equivalently, this adds the difference between the variant and wild-type embeddings to the stored wild-type embedding, so that a synonymous variant yields no change and the estimate is unaffected by scale offsets between embeddings. We then measured the change in predicted spatial expression, quantified as | log_2_ FC| between wild-type and mutant predictions averaged across tissue patches. Variant protein sequences were generated from the Ensembl GRCh38 coding sequence of each gene, and for each variant predictions were averaged over 50 spots sampled per tissue class. Because predictions are conditioned on local morphology, the estimate is tissue-conditioned and varies across the slide. We scored gnomAD-observed coding variants together with a sampled set of allele-frequency-absent variants, and for *BRCA1* and *BRCA2* additionally included all coding single-nucleotide variants curated in BRCA Exchange. Allele-frequency classes were defined from the gnomAD v4.1 whole-genome sequencing release as singleton (AF *<* 1 × 10^−5^), rare (1 × 10^−5^ ≤ AF *<* 1 × 10^−3^) and common (AF ≥ 1 × 10^−3^), with absent variants not observed in gnomAD. We evaluated nonsense versus missense variants, gnomAD allele-frequency classes [47], ClinVar clinical significance [48], COSMIC somatic variants in oncogenes and tumour suppressor genes [49], and ENIGMA-classified *BRCA1* /*BRCA2* variants [50]. Variants were evaluated in a single lung cancer sample (LC1) [14]. For the aggregate severity comparison (Fig. 6A), germline variants were assessed in regions manually annotated as normal and somatic variants in regions manually annotated as tumour by an expert pathologist, consistent with their respective tissue origins, whereas the spatial variant-effect maps (Fig. 6B) report the signed per-spot log_2_ FC across all annotated regions.

### 4.17 Language-guided spatial expression

For natural-language querying, gene queries were replaced with text embeddings of free-text descriptions of biological processes [27], which were passed to the cross-attention decoder to produce one spatial expression map per query. Performance was evaluated as the area under the precision-recall curve (AUPRC) against tissue annotations (normal versus tumour) and compared with CLIP [51] and the pathology-specific language model QUILT [52] using the same queries across the USZ lung squamous cell carcinoma, USZ clear cell renal cell carcinoma and WSSS4LUAD datasets.

### 4.18 Statistics and reproducibility

Predictive accuracy was reported as the gene-wise Pearson correlation between predicted and measured expression, computed and ranked as described in “Evaluation metric and ground-truth correction”. Results are shown as mean ± standard error across samples, genes or seeds as indicated. Sample-efficiency experiments were repeated across random seeds, and scaling behaviour was summarised with power-law fits (*R*^2^ reported). Group comparisons of marker expression used the two-sided Wilcoxon rank-sum test. No statistical method was used to predetermine sample size, and dataset splits were performed at the whole-slide-image or patient level, as appropriate, to avoid leakage between training and evaluation.

### 4.19 Implementation

DeepSpot-M is implemented in PyTorch and PyTorch Lightning.

## 5 Data availability

The TCGA virtual spatial transcriptomics atlas (28,664 whole-slide images across 32 cancer types) is available at https://huggingface.co/datasets/ratschlab/TCGA_virtual_spatial_transcriptomics_atlas. The virtual single-cell transcriptome-wide profiles inferred by DeepSpot-M for all 59 HEST-1K 10x Xenium samples, spanning 22 tissue types, are available at https://huggingface.co/datasets/ratschlab/HEST_Xenium_virtual_spatial_transcriptomics.

The public datasets analysed in this study are available from their original publications, HEST-1K [36], the lung 10x Xenium dataset [35], the mouse spatial total RNA-sequencing dataset [46], WSSS4LUAD [40] and TCGA (https://portal.gdc.cancer.gov). Malignant-versus-normal annotations for TCGA-BRCA were obtained from [39]. The TCGA tertiary lymphoid structure annotations were obtained from [57] (https://zenodo.org/records/10614928). The MOSAIC Window cohorts [28] are available from the European Genome-phenome Archive under accession EGAS50000000689 (https://ega-archive.org/studies/EGAS50000000689), subject to approval by the MOSAIC Data Access Committee. The USZ lung squamous cell carcinoma and clear cell renal cell carcinoma cohorts [14] are available from the original study under its access conditions. Germline allele frequencies were queried from the gnomAD v4.1 whole-genome sequencing (WGS) release [47] using the gnomAD database tool (https://github.com/KalinNonchev/gnomAD_DB).

## 6 Code availability

DeepSpot-M source code and trained model weights are available at https://github.com/ratschlab/ DeepSpotM.

## Acknowledgements

We thank the patients and their families and the clinical and laboratory teams supporting them, whose contributions made this study possible.

This study makes use of data generated by the MOSAIC consortium (Owkin, Charité – Universitätsmedizin Berlin, Lausanne University Hospital - CHUV, Erlangen Hospital, Gustave Roussy Institute, University of Pittsburgh) and made available through the MOSAIC Window initiative. Readers should note that the MOSAIC consortium bears no responsibility for the further analysis or interpretation of these data beyond what published by the MOSAIC consortium partners.

We also use data from the Tumor Profiler (TuPro) study [58] and thank the Tumor Profiler Consortium (https://tumorprofilercenter.ch).

Figures were created using a paid subscription to BioRender (https://www.biorender.com).

This work was supported as part of the Swiss AI Initiative (https://swiss-ai.org) by a grant from the Swiss National Supercomputing Centre (CSCS) under project ID a150 on Alps. Computational data analysis was performed at Leonhard Med (https://sis.id.ethz.ch/services/sensitiveresearchdata/), a secure trusted research environment at ETH Zurich.

## 7 Author contributions

Conceptualization and study design: K.N., V.H.K. and G.R. Model design and development: K.N. Experiments, analysis and visualization: K.N. Resources and data: K.S. and S.D. (USZ data and expert tertiary lymphoid structure annotations). Supervision and funding: V.H.K. and G.R. Manuscript drafting: K.N., V.H.K. and G.R. Manuscript review: all authors critically reviewed the manuscript.

