## Supplementary information for "DeepSpot-M: a multimodal foundation model for transcriptome-wide virtual spatial transcriptomics from histology"

### Supplementary Information: DeepSpot-M

#### Supplementary Methods

##### Specialised histology-to-spatial-transcriptomics baselines

For the MOSAIC Window benchmark, DeepSpot-M was compared with four specialised histology-to-spatial-transcriptomics methods trained separately on each target cohort. These methods were chosen to represent the main families of supervised spatial expression prediction from H&E, spanning direct convolutional regression, contrastive image-expression retrieval, spatial-context modelling and multiscale morphology integration. In contrast to DeepSpot-M, all specialised baselines use cohort-specific training and fixed output gene sets; they therefore cannot directly query genes outside the panel used for training. Performance was evaluated with the same gene-wise Pearson correlation metric used in the main text and summarised across top-$K$ gene sets.

***ST-Net.***

ST-Net [1] is a supervised convolutional model that predicts spatial transcriptomics measurements from H&E image patches. In our benchmark, ST-Net used DenseNet-121 image features as its feature extractor, following its official implementation (https://github.com/bryanhe/ST-Net), trained on fixed gene targets; it was introduced for linking breast tumour morphology with spatial gene expression.

***BLEEP.***

BLEEP [2] (Bi-modaL Embedding for Expression Prediction) learns a joint image-expression embedding with contrastive learning. In our benchmark, BLEEP used ResNet-50 image features as its feature extractor, following its official implementation (https://github.com/bowang-lab/BLEEP). At inference, expression for a query histology patch is imputed from expression profiles in the reference set through this learned shared space.

***DeepSpot.***

DeepSpot [3] predicts spatial transcriptomics from H&E images by combining pathology foundation-model features with a deep-set architecture over sub-tiles and neighbouring spots, thereby incorporating local morphology and spatial context. In our benchmark, DeepSpot used frozen Midnight [4] embeddings as its feature extractor, the same vision backbone as DeepSpot-M. This departs from the original DeepSpot, which selected the best-performing pathology foundation model per cancer type among UNI, Phikon and H-optimus-0. We instead fixed Midnight, a more recent state-of-the-art pathology foundation model [4], as the vision backbone across both methods, so that performance differences reflect the model architecture rather than the choice of feature extractor. Code: https://github.com/ratschlab/DeepSpot.

***MISO.***

MISO [5] is a multiscale integration model for spatial omics and tumour morphology. It extracts tile-level and patch-level pathology features, incorporates neighbouring spots and supports super-resolved expression inference through model distillation. In our benchmark, MISO used H-optimus-0 mini [6] as its feature extractor, following its official implementation. Code: https://github.com/owkin/miso_code.

#### Histology-to-bulk-transcriptome baseline

***Sequoia.***

For the pseudo-bulk reconstruction benchmark, DeepSpot-M was additionally compared with Sequoia [7], a specialised histology-to-bulk-transcriptome model for digital profiling of cancer transcriptomes from whole-slide images. Sequoia predicts bulk RNA-seq expression directly from H&E using grouped vision attention. We used the pretrained Sequoia weights provided by the authors on Hugging Face under https://huggingface.co/gevaertlab, using split 0 for each selected model. When an exact cancer-type model was unavailable for a MOSAIC Window cohort, we selected the closest available TCGA cancer model: urothelial carcinoma was evaluated with BLCA, glioblastoma with GBM, mesothelioma with LUAD, ovarian cancer with BRCA and diffuse large B-cell lymphoma with SKCM. For each sample, UNI image features were clustered into 100 centroids before Sequoia inference, and predictions were evaluated only for genes shared with the MOSAIC bulk RNA-seq table. In our benchmark, Sequoia represents a bulk-specific upper comparator for DeepSpot-M, which is trained only on spatial transcriptomics and evaluated after aggregating predicted spot-level expression into pseudo-bulk profiles. Code: https://github.com/gevaertlab/sequoia-pub.

#### Language-guided spatial-query baselines

For the natural-language spatial-query benchmark, DeepSpot-M was compared with two image-text representation models applied to the same H&E image regions and free-text biological queries. We used OpenAI CLIP ViT-B/32 [8] from https://huggingface.co/openai/clip-vit-base-patch32 and QUILT/QuiltNet-B-32 [9], a pathology-specific vision-language model, from https://huggingface.co/wisdomik/QuiltNet-B-32. For each query, baseline scores were computed from the similarity between the query text embedding and the corresponding image-region embedding, and performance was evaluated against the same tissue annotations as DeepSpot-M using the area under the precision-recall curve.

### Extended Data Figures

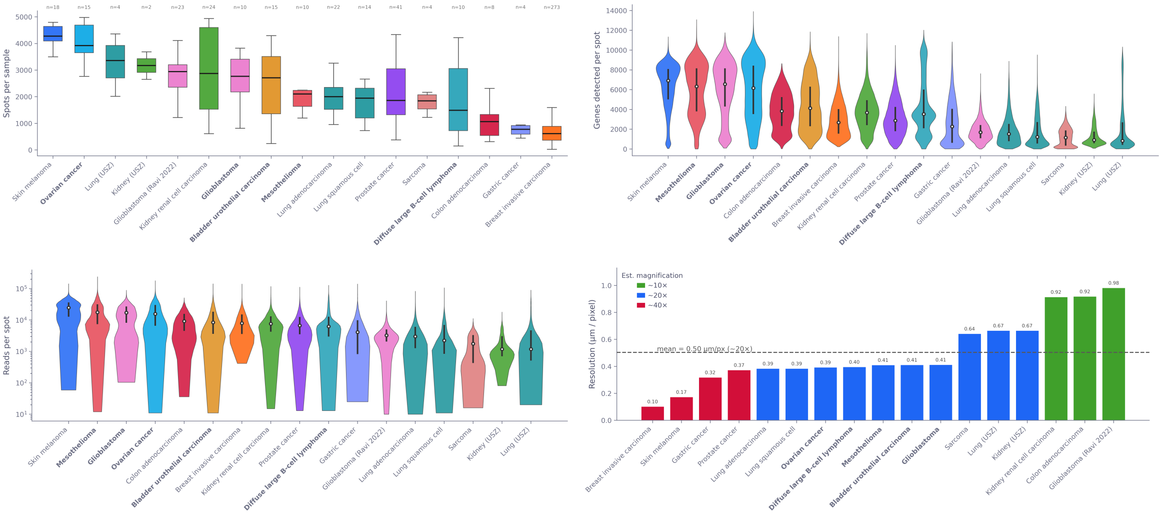


**Extended Data Fig. 1 Characteristics of the training corpus.**
**a**, Number of 10x Visium spots in each of the 15 training datasets, together comprising the 730,000 paired profiles used for training. **b**, Number of genes detected per dataset after preprocessing, relative to the harmonised vocabulary of 19,338 protein-coding genes. A binary gene mask ensures that loss and metrics are computed only on genes measured in each dataset. **c**, Distribution of per-spot sequencing depth across datasets, illustrating the technical variability in capture efficiency across cohorts and institutions that the unified preprocessing workflow harmonises. **d**, H&E whole-slide scanning magnification per dataset; tiles are extracted at a consistent physical resolution to control for differences in native scanning magnification.

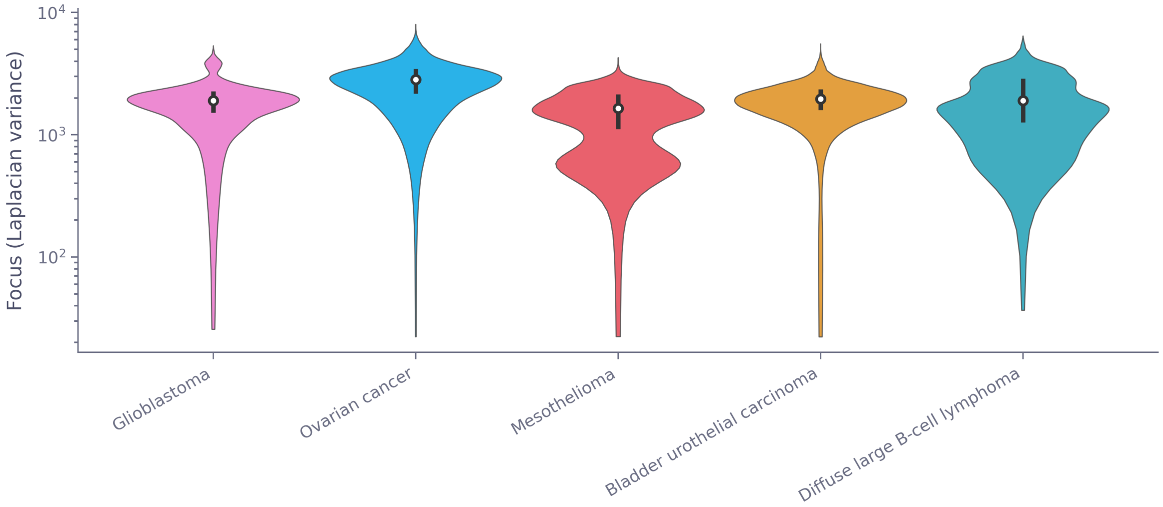


**Extended Data Fig. 2 Image focus across MOSAIC Window benchmark slides.**
Per-cohort image focus quality for the five MOSAIC Window benchmark datasets. The mesothelioma cohort shows reduced focus quality and the fewest available spots (Extended Data Fig. 1), consistent with its lower absolute prediction accuracy despite a preserved ranking of methods.

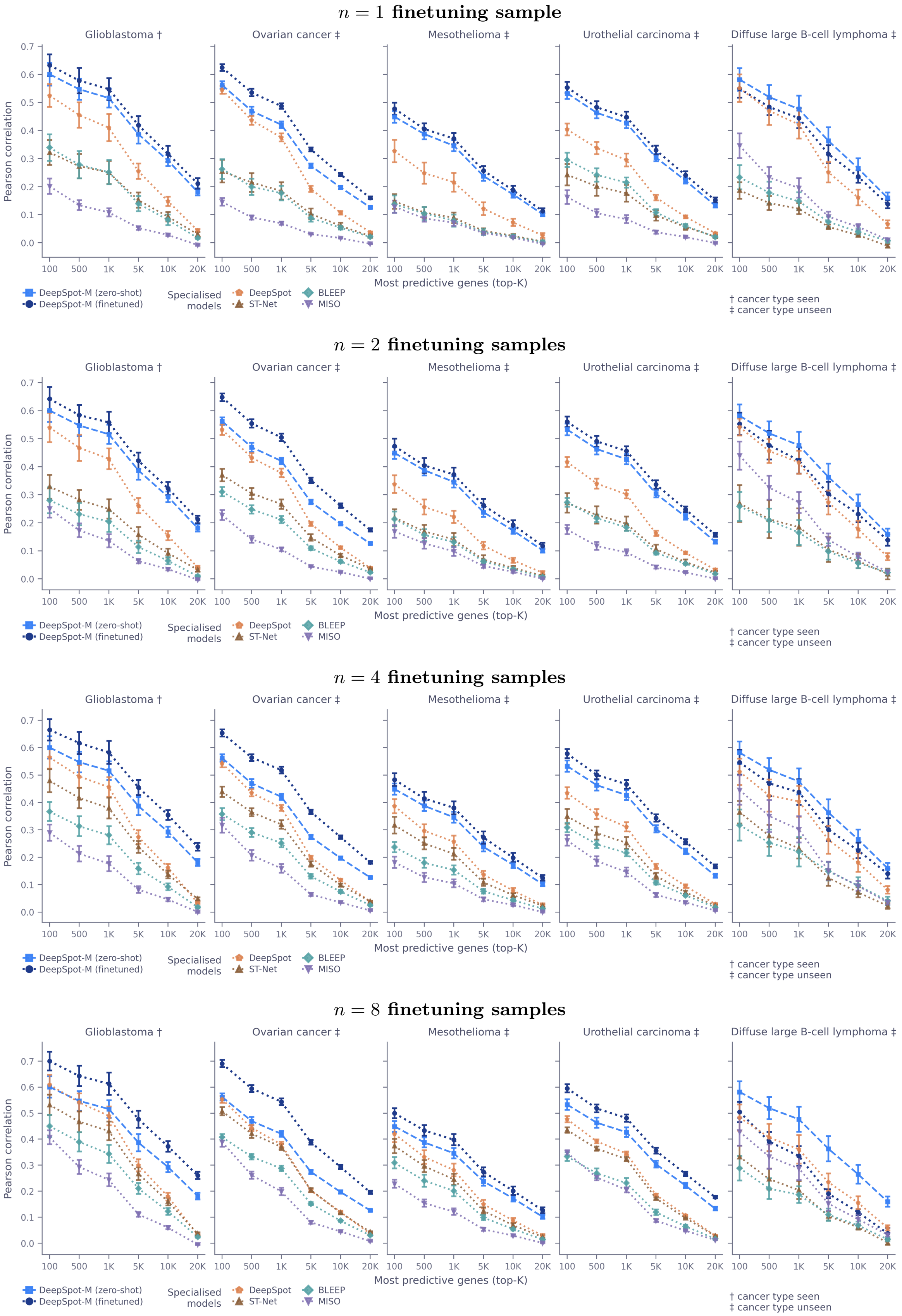


**Extended Data Fig. 3 Benchmark consistency across finetuning sample sizes.** Top-$K$ gene-wise Pearson correlation on the five MOSAIC Window benchmark cohorts (as in Fig. 2A), with DeepSpot-M finetuned on $n = 1$, 2, 4 and 8 cohort-specific samples (top to bottom, labelled above each row). DeepSpot-M outperforms the specialised baselines across all settings, showing that its advantage is consistent across the number of cohort-specific samples used for finetuning.

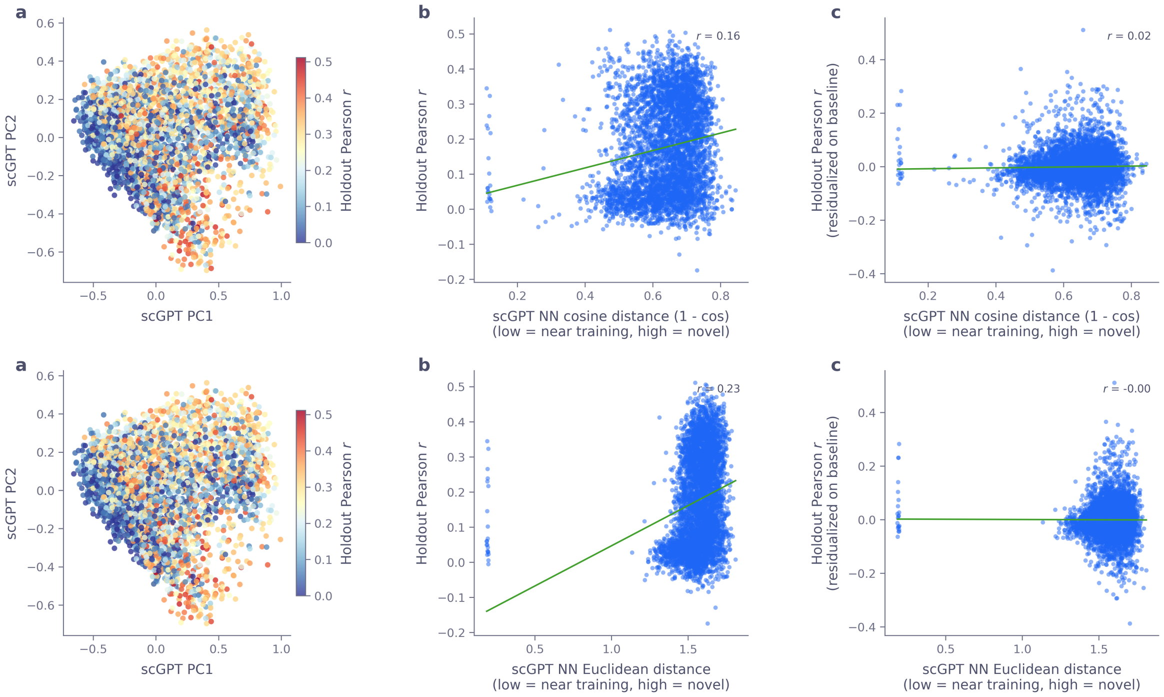


**Extended Data Fig. 4  Holdout-gene accuracy is not explained by nearest-neighbour distance.** Per-gene zero-shot Pearson correlation for held-out genes as a function of their distance to the nearest training gene in scGPT embedding space, using cosine (top) and euclidean (bottom) distance. Prediction accuracy is not explained by proximity to the nearest training gene, indicating that DeepSpot-M exploits broader gene-gene structure rather than copying the closest neighbour.

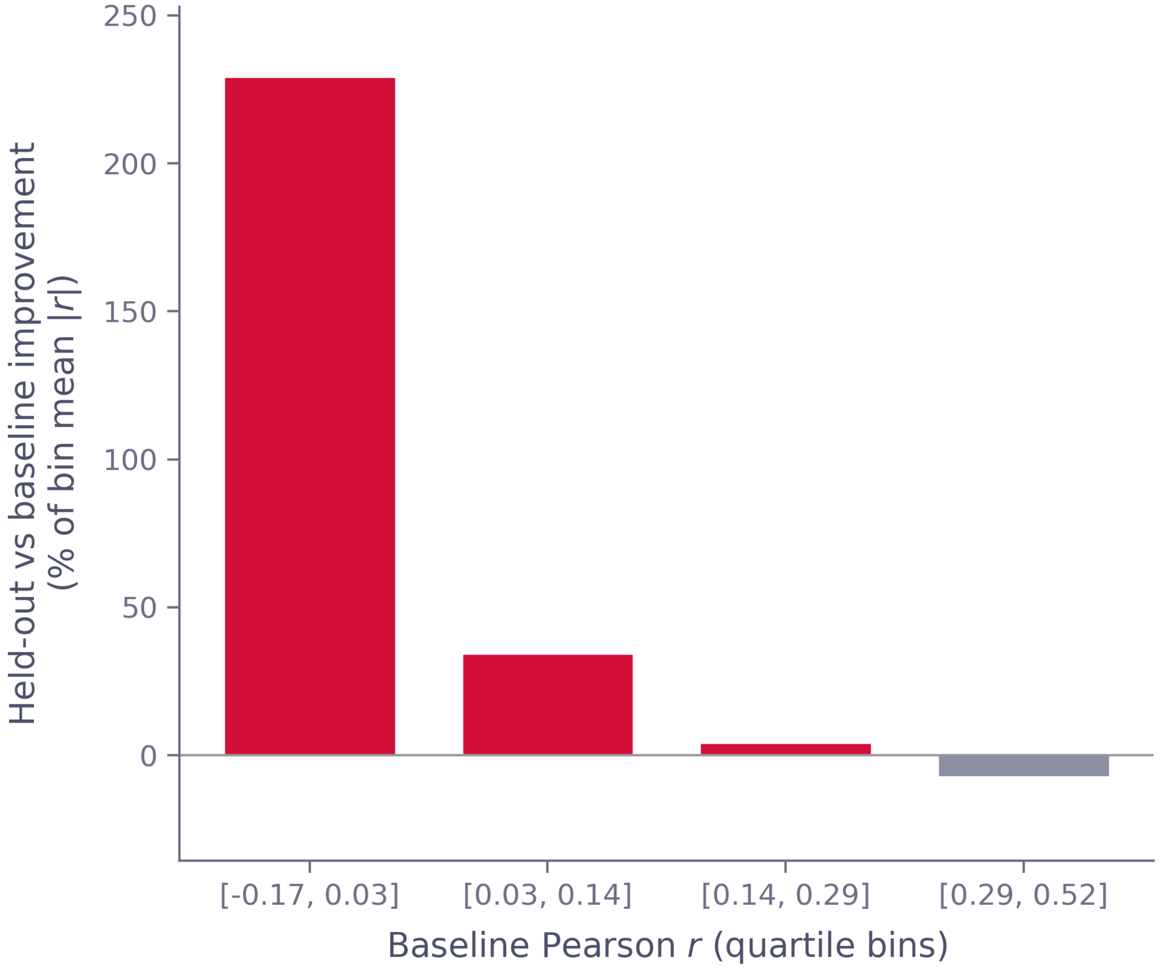


**Extended Data Fig. 5 Gene-holdout versus full model by gene predictability.**
Per-gene difference in Pearson correlation between the gene-holdout model and the full model trained on all genes, binned by gene predictability. The full model is more accurate for the most predictable genes, whereas the gene-holdout model performs better for less predictable genes, consistent with the holdout model relying more strongly on gene-gene structure encoded in the biological embeddings.

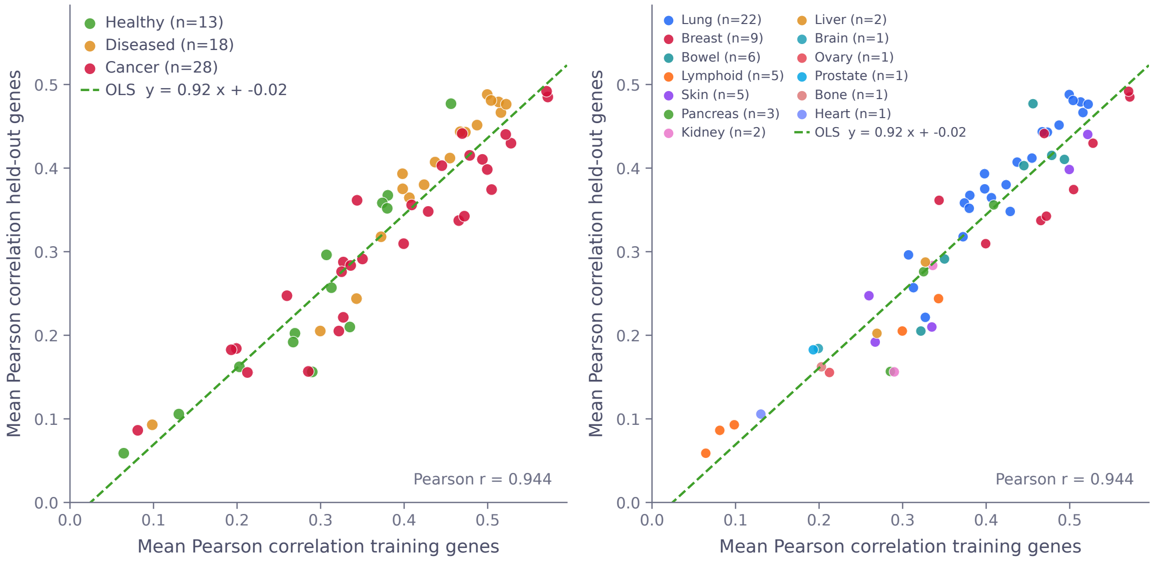


**Extended Data Fig. 6 Held-out-gene accuracy tracks measured-panel accuracy across HEST-1K Xenium samples.**
Per-sample mean Pearson correlation on held-out genes versus measured-panel genes across the 59 HEST-1K 10x Xenium samples [10], spanning 22 tissue types, coloured by **a**, disease and **b**, organ system. Samples with higher measured-panel accuracy also tend to have higher held-out-gene accuracy (Pearson $r = 0.94$, $n = 22$ tissues).

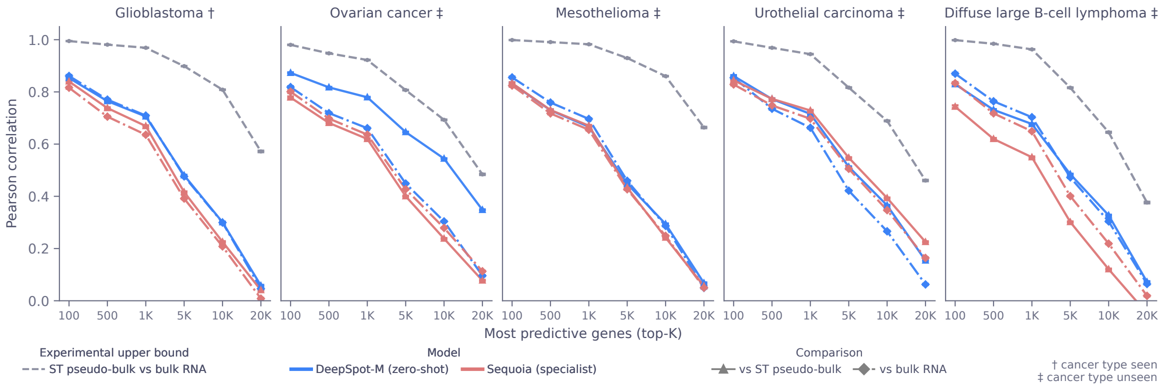


**Extended Data Fig. 7 DeepSpot-M reconstructs bulk transcriptomes from histology.**
Bulk benchmark of DeepSpot-M (zero-shot) and Sequoia [7], a specialised histology-to-bulk model, on the five MOSAIC Window cohorts. For each sample, predicted spatial expression is aggregated into a pseudo-bulk profile and correlated gene by gene with matched bulk RNA-seq across the top-predictive genes (N). Although DeepSpot-M is not trained on bulk RNA-seq, it matches or exceeds Sequoia across cohorts and approaches the experimental upper bound (grey dashed line, measured ST pseudo-bulk versus bulk RNA-seq).

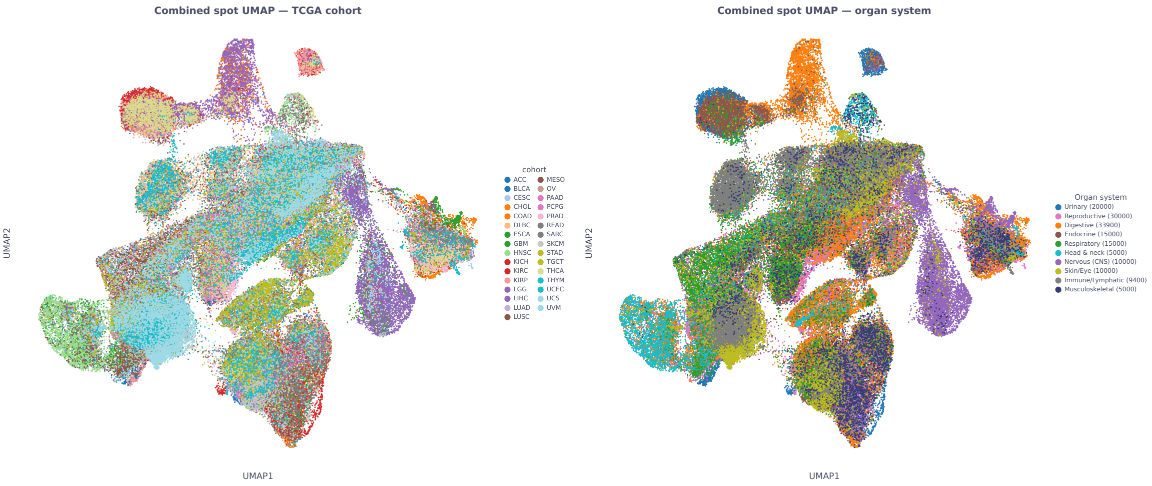


**Extended Data Fig. 8 Pan-cancer UMAP of the TCGA virtual atlas.**
UMAP of DeepSpot-M predicted spatial expression across the TCGA virtual atlas, with each spot coloured by **a**, TCGA cancer-type cohort and **b**, organ system. Spots from the same cancer type form coherent regions, and cancers from related organ systems occupy neighbouring regions, indicating that the predicted profiles retain cancer-type- and organ-specific structure.

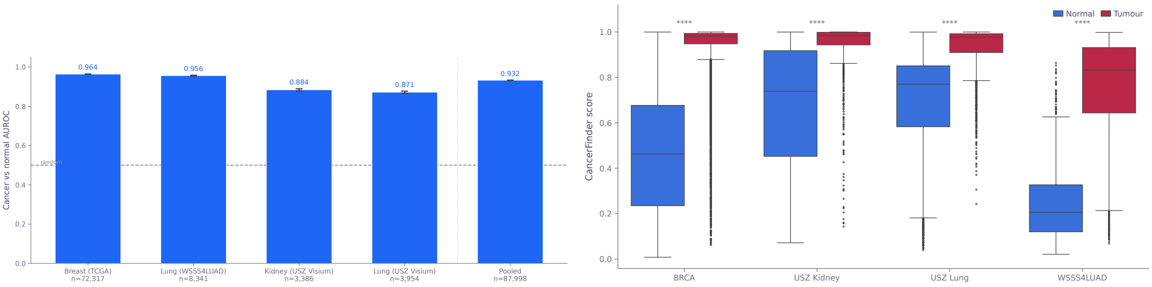


**Extended Data Fig. 9 Predicted malignancy scores agree with expert annotations.**
CancerFinder [11] applied to DeepSpot-M predicted expression and evaluated against expert malignant-versus-normal annotations across four datasets: TCGA-BRCA breast cancer [12], WSSS4LUAD lung [13], and the USZ Visium lung and kidney cohorts [3]. **a**, Cancer-versus-normal classification AUROC. DeepSpot-M distinguishes malignant from non-malignant tissue with high accuracy across all datasets (AUROC 0.87-0.96 per dataset, 0.93 pooled across $n = 87{,}998$ annotated regions), well above the random baseline (0.5, dashed line). Error bars indicate bootstrap confidence intervals. **b**, Distribution of malignancy scores in malignant versus normal regions. Scores are significantly higher in malignant regions in every dataset (Wilcoxon rank-sum test, $P < 0.001$).

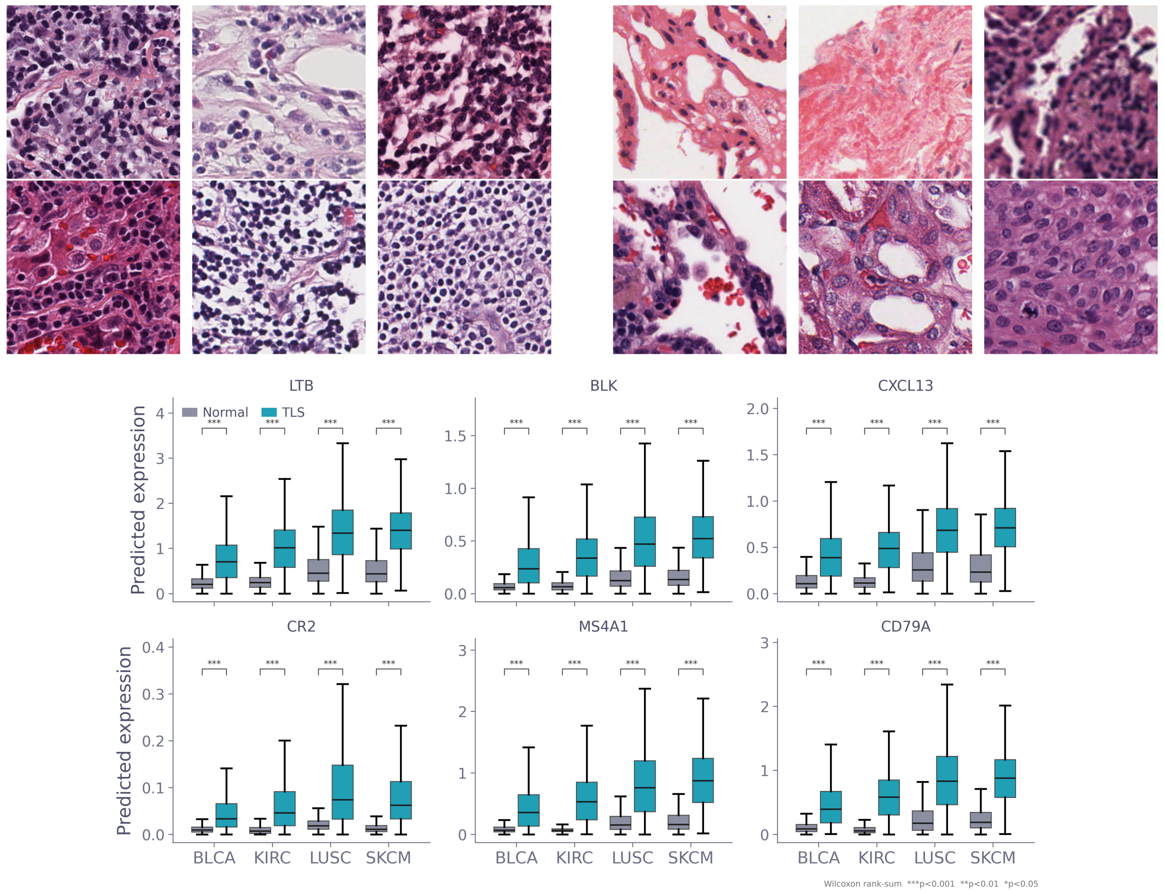


**Extended Data Fig. 10 Predicted TLS-marker expression in TLS versus non-TLS regions of the TCGA atlas.**
Top, example H&E patches of tertiary lymphoid structure (TLS) and normal tissue. Bottom, predicted expression of TLS and B-cell markers (*LTB*, *BLK*, *CXCL13*, *CR2*, *MS4A1* and *CD79A*) in TLS versus normal regions across BLCA, KIRC, LUSC and SKCM. All markers are significantly higher in TLS (Wilcoxon rank-sum test, $P < 0.001$).

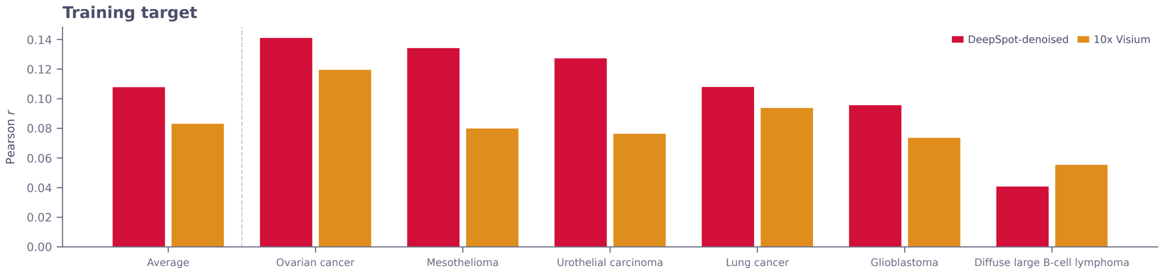


**Extended Data Fig. 11 Training on DeepSpot-denoised targets improves prediction accuracy.**
Mean Pearson $r$ for DeepSpot-M trained on DeepSpot-denoised expression targets (red) versus raw 10x Visium measurements (orange). DeepSpot-denoised targets, which reduce dropout noise through spatial smoothing, consistently improve performance across all held-out cancer types. The average improvement is $\sim$0.023 Pearson $r$, confirming that self-distillation from denoised targets provides a stronger training signal for spatial gene expression prediction.

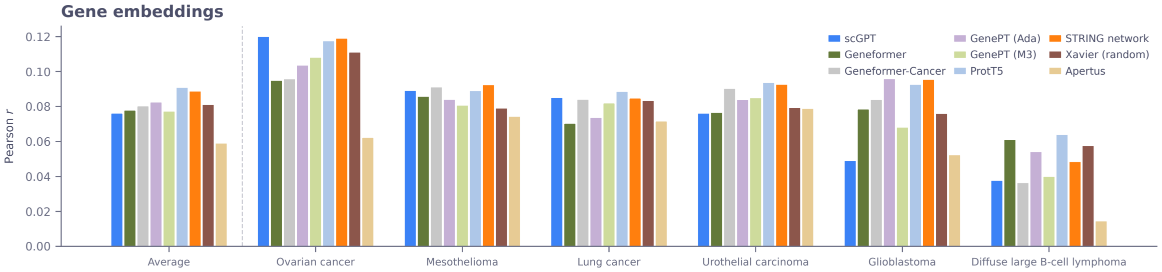


**Extended Data Fig. 12 Gene embedding comparison across seven biological sources and a random baseline.**
Mean Pearson $r$ for DeepSpot-M using different pretrained gene embeddings as query initialisations and router inputs. The seven biological sources are scGPT [14] (512-d), Geneformer [15] (768-d), Geneformer-Cancer [15] (768-d, cancer-finetuned), GenePT-Ada [16] (1536-d), GenePT-M3 [16] (3072-d), ProtT5 [17] (1024-d) and STRING network [18] (512-d), benchmarked against a Xavier random initialisation.

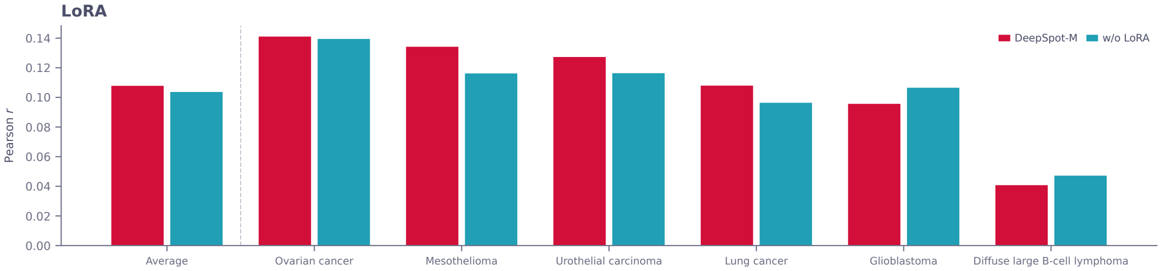


**Extended Data Fig. 13 LoRA adaptation of the vision encoder improves performance.**
Mean Pearson $r$ for DeepSpot-M with (red) and without (teal) LoRA adapters on the vision encoder. LoRA ($r = 16$, $\alpha = 32$) adds $< 0.5\%$ trainable parameters to the frozen Midnight [4].

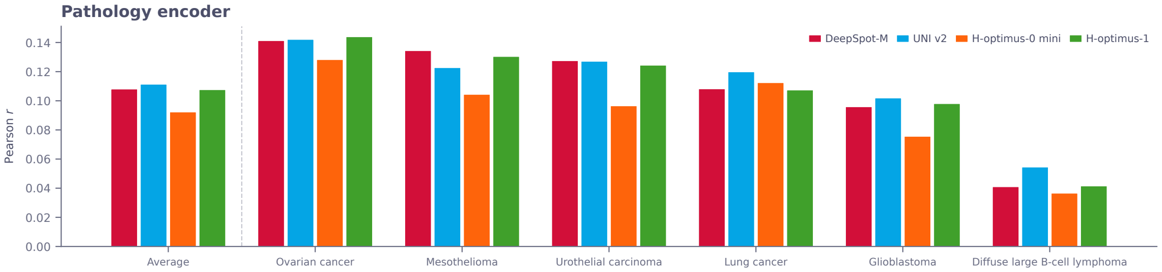


**Extended Data Fig. 14 Comparison of pathology foundation models as vision encoders.**
Mean Pearson $r$ for DeepSpot-M using different pathology foundation models, namely Midnight [4] (ViT-Giant, default), UNI v2 [19] (ViT-Large), H-optimus-0 mini [6] and H-optimus-1 [20]. All encoders achieve comparable average performance, with UNI v2 and H-optimus-1 slightly outperforming Midnight on several cancer types. This indicates that DeepSpot-M's cross-attention architecture generalises across vision backbones and can benefit from future improvements in pathology foundation models.

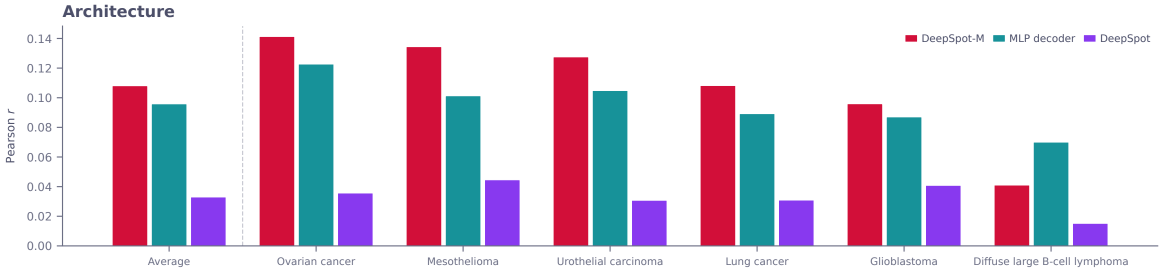


**Extended Data Fig. 15 DeepSpot-M outperforms prior architectures in leave-one-dataset-out evaluation.**
Mean Pearson $r$ across 19,338 genes for DeepSpot-M (cross-attention gene decoder with gene router), an MLP decoder baseline (shared representation, no cross-attention), and DeepSpot [3] (deep-set neural network). Results are shown per held-out cancer type and as a cross-cancer average. DeepSpot-M consistently outperforms both baselines, with the largest gains on mesothelioma and urothelial carcinoma. DeepSpot, which compresses all genes into a single shared feature vector, achieves markedly lower performance, confirming that per-gene cross-attention is essential for transcriptome-wide prediction.

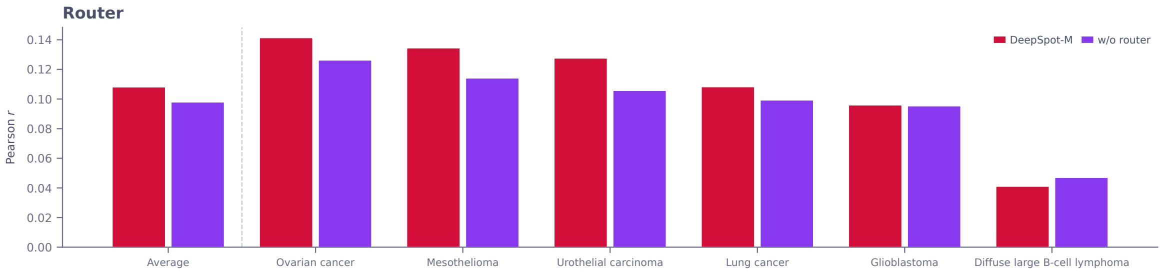


**Extended Data Fig. 16 The gene router hypernetwork improves prediction accuracy.**
Mean Pearson $r$ for DeepSpot-M with (red) and without (purple) the gene router hypernetwork. The router, which generates per-gene output projections from biological embeddings, provides consistent improvements across most cancer types, with the largest gains on ovarian cancer and mesothelioma. In addition to improving supervised prediction, the router enables zero-shot prediction of genes not seen during training.

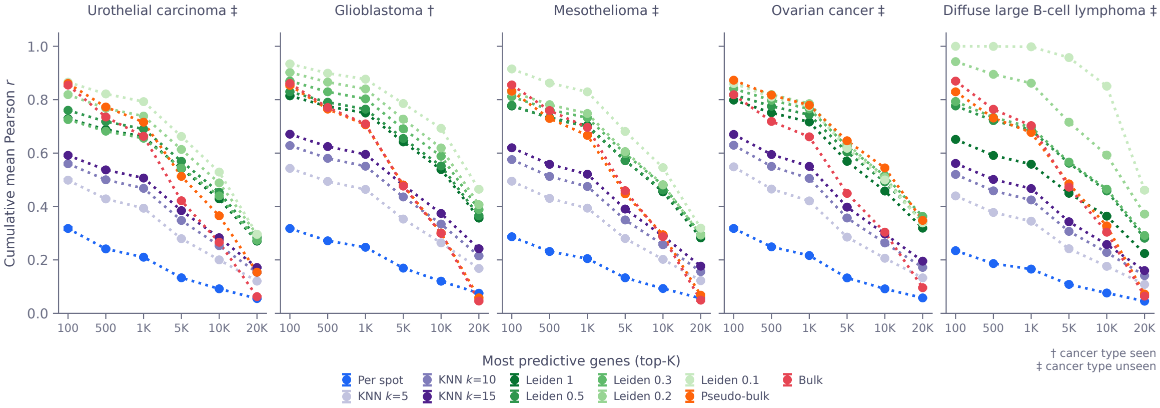


**Extended Data Fig. 17 Spot-level Visium ground-truth correction ablations.** Cumulative mean gene-wise Pearson correlation across the top-predictive genes for five MOSAIC Window cohorts. Per-spot evaluation compares DeepSpot-M predictions directly with measured 10x Visium expression. KNN curves correct the noisy Visium reference by averaging measured expression over neighbourhoods in a PCA latent space of log-normalised measured ST expression using $k = 5, 10$ or 15 neighbours, with $k = 5$ the default used for all reported correlations. Leiden curves analogously correct the measured reference by averaging expression within Leiden clusters at resolutions 1, 0.5, 0.3, 0.2 and 0.1. Pseudo-bulk averages expression over all spots in a sample, and bulk compares with matched bulk RNA-seq. These ablations test whether molecular-latent-space smoothing or cluster-level averaging mitigates 10x Visium sparsity, dropout and spot-level noise in the evaluation reference [21, 22].

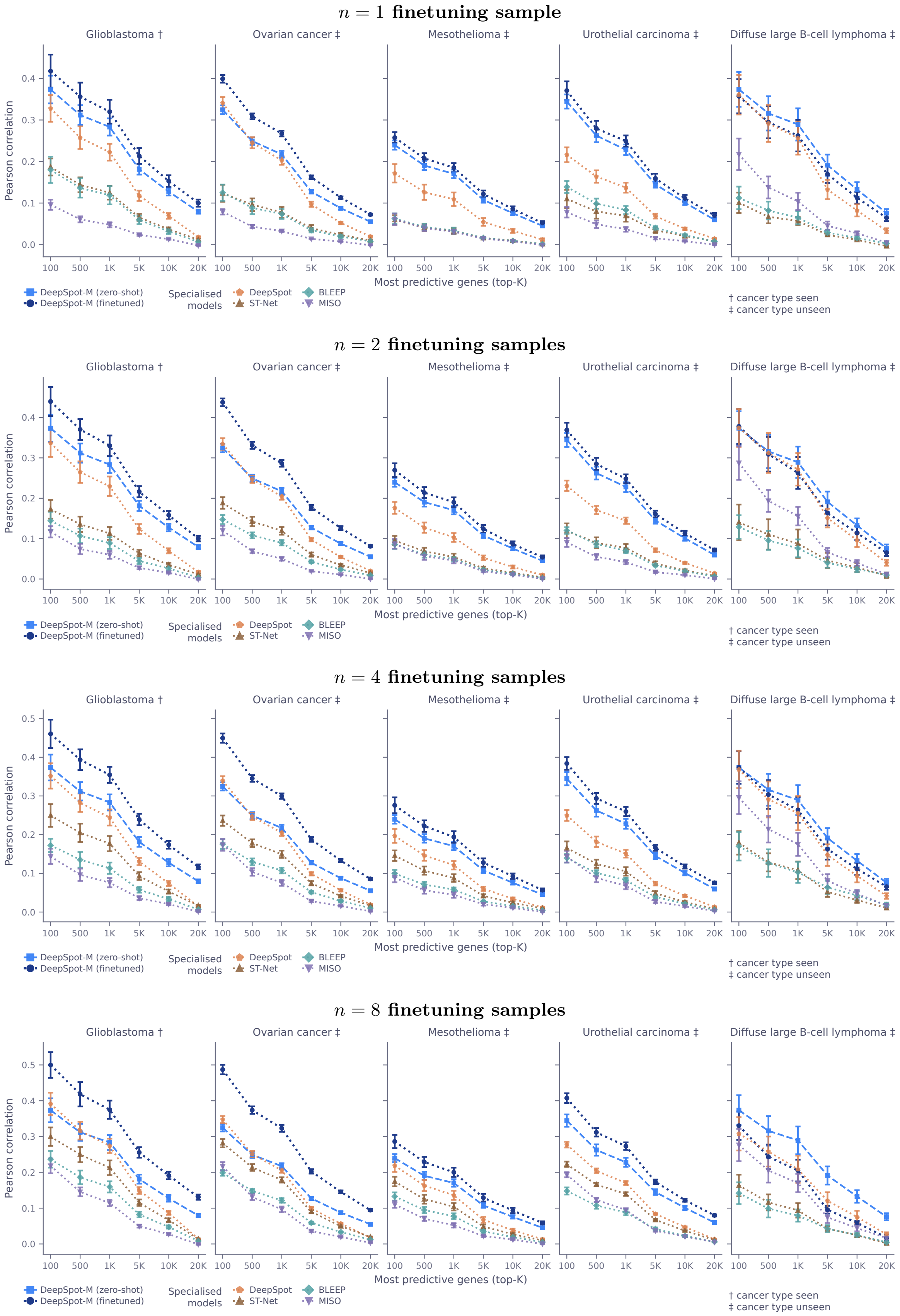


**Extended Data Fig. 18  Benchmark consistency on raw measured counts.**
Top-$K$ gene-wise Pearson correlation on the five MOSAIC Window benchmark cohorts (as in Fig. 2A and Extended Data Fig. 3), with correlations computed directly against raw 10x Visium counts rather than the neighbourhood-smoothed reference used in the main analysis. DeepSpot-M finetuned on $n = 1$, 2, 4 and 8 cohort-specific samples (top to bottom, labelled above each row) outperforms the specialised baselines across all settings, showing that its advantage persists without ground-truth smoothing.
